# A simple model of COVID-19 explains disease severity and the effect of treatments

**DOI:** 10.1101/2021.11.29.21267028

**Authors:** Steven Sanche, Tyler Cassidy, Pinghan Chu, Alan S. Perelson, Ruy M. Ribeiro, Ruian Ke

## Abstract

Considerable effort was made to better understand why some people suffer from severe COVID-19 while others remain asymptomatic. This has led to important clinical findings; people with severe COVID-19 generally experience persistently high levels of inflammation, slower viral load decay, display a dysregulated type-I interferon response, have less active natural killer cells and increased levels of neutrophil extracellular traps. How these findings are connected to the pathogenesis of COVID-19 remains unclear. We propose a mathematical model that sheds light on this issue. The model focuses on cells that trigger inflammation through molecular patterns: infected cells carrying pathogen-associated molecular patterns (PAMPs) and damaged cells producing damage-associated molecular patterns (DAMPs). The former signals the presence of pathogens while the latter signals danger such as hypoxia or the lack of nutrients. Analyses show that SARS-CoV-2 infections can lead to a self-perpetuating feedback loop between DAMP expressing cells and inflammation. It identifies the inability to quickly clear PAMPs and DAMPs as the main contributor to hyperinflammation. The model explains clinical findings and the conditional impact of treatments on disease severity. The simplicity of the model and its high level of consistency with clinical findings motivate its use for the formulation of new treatment strategies.

## INTRODUCTION

COVID-19 symptoms severity differs wildly between infected individuals. Some individuals are infected without experiencing many of the characteristic symptoms, such as fever, coughs, body aches and the loss of taste or smell.(1) At the other end of the spectrum, a substantial minority will experience more extreme symptoms, such as acute respiratory distress syndrome and thrombotic complications that can lead to organ failure and death.(1,2) What distinguishes individuals experiencing more severe symptoms has been extensively studied.(3) These studies have identified a set of risk factors associated with severe COVID-19 such as older age, obesity, diabetes and past or present cancer.(3) COVID-19 severity likely depends on both the trajectory of the viral infection and the trajectory of the inflammatory response. An association was found between endothelial cell expression of angiotensin-converting enzyme 2 (ACE2), the receptor for severe acute respiratory syndrome coronavirus-2 (SARS-CoV-2) entry to host cells, and the presence of microthrombi in major organs such as the lung, heart, brain, and liver.(4,5) This suggests that the spread of the infection may be responsible for damage to vital tissues and organ dysfunction. It also underlines the necessity of an appropriate innate and adaptive immune response to limit the spread of the infection. To this effect, inflammation plays a crucial role by coordinating the immune response. However, elevated inflammatory markers in COVID-19 patients (IL-1*β*, IL-2R, IL-6, IL-8, IL-10, TNF-*α*, to name a few) have been associated with severity of symptoms, the need for ventilation and deaths.(6–8) This suggests that a sustained or exaggerated inflammatory response (hyperinflammation) may play an important role in determining disease severity. In particular, an inappropriate innate immune response was pointed out as a significant contributor to the hyperinflammatory state in COVID-19.(9,10) Despite the large effort provided by the scientific community, there are still many unknowns with regards to the mechanistic drivers of severe COVID-19. In turn, this knowledge gap hampers our ability to find new treatment strategies aiming to improve clinical outcomes.

Our main objective was to provide a simple quantitative framework to understand the pathogenesis of severe COVID-19 and to determine the importance of potential mechanisms. We aimed to provide a model that is simple and yet adequately captures the main clinical findings. Many within-host models of SARS-CoV-2 infection have been published (11–17) with a wide range of model complexities (from less than 10 to more than 80 parameters). These models mostly differ in terms of the complex interactions between the viral infection and the immune response that they include in their formulation (see Discussion for details). The large difference between model formulations suggests that identifying the key elements having an impact on clinical outcomes is a difficult task.

The model we formulated focuses on cells that trigger inflammation through molecular patterns: infected cells carrying pathogen-associated molecular patterns (PAMPs) and damaged cells producing damage-associated molecular patterns (DAMPs). We show that the clearance rate of infected and damage cells by the innate immune response is of the utmost importance to reach a state of resolved of inflammation. Despite its simplicity, our model can explain the following findings: i) severe COVID-19 tends to be accompanied by hyperinflammation, ii) those with severe COVID-19 generally experience a similar viral trajectory as mild cases, albeit with a slower viral load decay after the peak, iii) the complex and conditional effect of antivirals and corticosteroids on disease severity, iv) an inefficient type-I IFN response is associated with severe COVID-19, and v) generation of bystander cell damage and infective removal of these cells are a critical component of severity. Note that this last point is reminiscent of clinical observations that, for example, less cytotoxic NK cells and higher levels of neutrophil extracellular traps (NETs) are associated with severe COVID-19. Overall, the simplicity of the model we propose along with its high level of consistency with clinical observations suggest it is an adequate framework for the study of COVID-19 pathogenesis and the effect of therapy.

## METHODS

### The Model

Our goal was to formulate a model that is simple enough to guide intuition, yet complex enough to allow relating with clinical outcomes. A schematic representation is provided in Fig. 1. The model is described below.

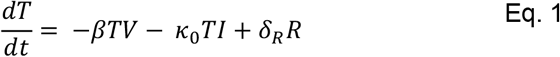

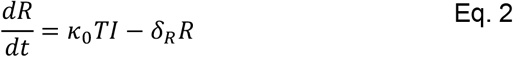

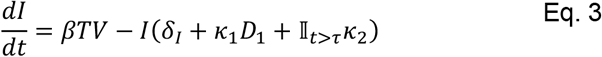

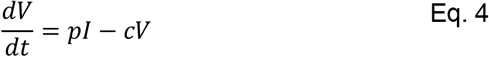

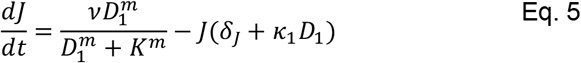

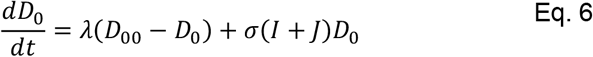

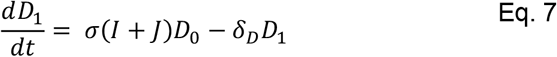

**Figure 1.**
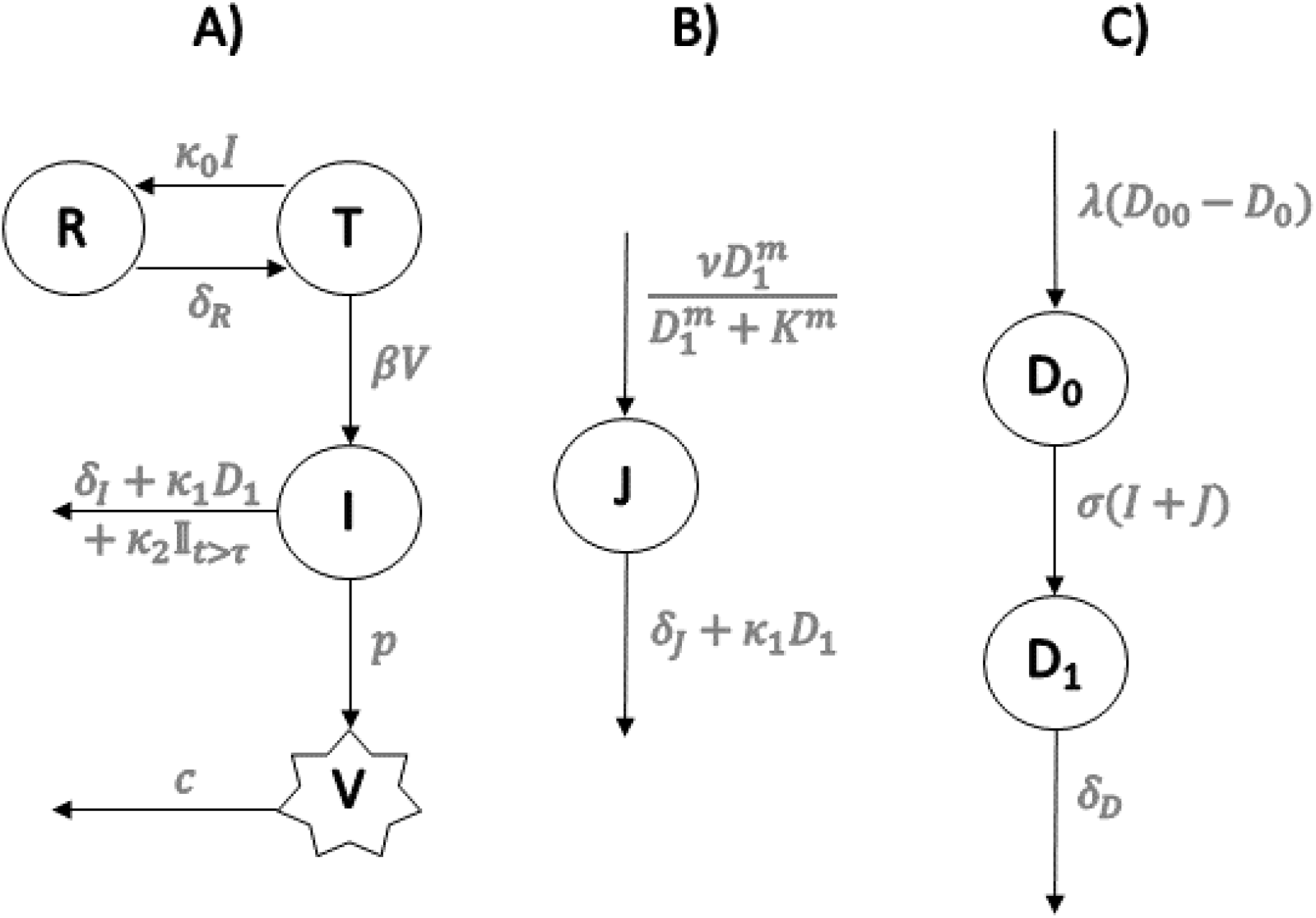
Schematic representation of the model described by Eqs. 1-7. Target cells (*T*) transition to a productively infected state (*I*) after successful infection by virions (*V*) at rate β. Virions are cleared at per capita rate c, while new virions are produced by infected cells at rate *p*. Target cells become refractory to infection (*R*) at rate *κ*_0_*I* (we assume target cells are exposed to a concentration of type-I IFN that is proportional to *I* and that puts the cells into an antiviral state). Resting innate immune cells (*D*_0_) become activated (*D*_1_) at rate σ(I+J), where *I* and *J* are the number of infected and damaged bystander cells, respectively (we assume the extent of PAMP and DAMP signaling is proportional to *I*+*J*). Also, activated immune cells (*D*_1_) die at per capita rate *δ*_*D*_. Damaged bystander cells (*J*) are generated from an extensive proinflammatory response at a rate that is a Hill function of the number of activated immune cells *D*_1_. Infected cells die due to viral cytopathic effects at rate *δ*_*I*_ and damaged cells die from their injury at rate *δ*_*J*_. The clearance of these cells also occurs by the action of activated innate immune cells at rate *κ*_1_*D*_1_. The effect of the adaptive immune response is modeled by adding a constant term κ_2_ to the clearance of infected cells at time τ after infection. Finally, homeostatic processes allow replenishment of the population of resting cells (*D*_0_) at rate λ(*D*_00_ − *D*_0_), where *D*_00_ is their homeostatic level.

#### Modeling the course of infection in absence of immune responses

We describe the course of the infection in the absence of immune response through cells susceptible to viral infection (*T*) that can become infected (*I*) by contact with virions (*V*). We assumed this process follows mass action with rate *βV*. Infected cells die at constant rate *δ*_*I*_ due to viral cytopathic effects. Infected cells produce virions at rate *p*, which are cleared at per capita rate *c*.

#### Modeling inflammation and the innate immune response

The model focuses on molecular patterns that initiate an inflammatory response. Pathogen associated molecular patterns (PAMPs) trigger an inflammatory response by signaling the presence of pathogens.(18) An inflammatory response can also be initiated in the absence of pathogenic infection through recognition of damage associated molecular patterns (DAMPs).(19) Various conditions promote DAMP expression including hypoxia, low levels of glucose and amino acids, exposure to heat, physical stress or exposure to toxic molecular products.(19–22) Many of the same conditions that promote DAMP expression can be induced by inflammation itself. For example, an excessive presence of neutrophil extracellular traps (NETs) released during inflammation is linked to immunothrombosis.(23) This can contribute to hypoxia and nutrient deprivation that can lead to DAMP expression.(24) Overall, a positive feedback loop between inflammation and DAMP expression may be triggered. (22,24) Interestingly, patients with severe COVID-19 have increased levels of NETs.(25,26) NETs and platelet dysregulation may be specific to COVID-19 and both are associated with lung microthrombi, conditions that can ultimately favor an inflammatory response in the lungs.(27)

The inflammatory response signals a need to eliminate infected or damaged cells and protect the rest of the organism from the perceived danger. Infected cells carrying PAMPs and uninfected cells producing DAMPs are represented by *I* and *J* in the model, respectively. In the organism, these molecular patterns promote the downstream recruitment and activation of numerous innate immune cells such as neutrophils, monocytes, macrophages and natural killer (NK) cells.(28) For simplicity, we lumped all cells that take part directly or indirectly (through cytokine signaling) in cytolytic or phagocytic activities into two model compartments, *D*_0_ and *D*_1_ representing resting and activated phenotypes, respectively. In our model, the rate at which *D*_0_ cells become *D*_1_ is proportional to the amount of PAMPs and DAMPs, themselves assumed proportional to the number of cells carrying these molecules, i.e. *σ*(*I* + *J*). We further assumed resting cells *D*_0_ have a homeostatic level *D*_00_, that they maintain through the recruitment of resting immune cells over the course of infection at rate *λ*. We assumed *D*_0_ cells are long-lived compared to the duration of acute infection, and that activated cells decay at rate *δ*_*D*_. The role of *D*_1_ in our model is to promote cytolytic and phagocytic activities to rid the system of PAMPs and DAMPs. Due to the innate nature of the modeled response, we assumed the effect to be similar on the decay of both *I* and *J* at per capita rate *κ*_1_*D*_1_. This was partly motivated by the behavior of NK cells, which target injured epithelial cells through stimulation of receptor NKG2D, as well as infected cells having downregulated major histocompatibility complex class 1 (MHC-I) molecules or upregulated MHC class I polypeptide–related sequence A (MICA) or sequence B (MICB).(29,30)

We assumed that high levels of inflammation can induce significant cellular stress and promote the expression of DAMPs in bystander cells. This is represented by the Hill term in Eq. 5 for the rate of generation of uninfected cells expressing DAMPs with *v* being the maximum damage rate, *K* being the *D*_1_ concentration leading to half of the maximum damage rate, and *m* is the Hill coefficient. We assumed damaged cells decay at rate *δ*_*J*_ < *δ*_*I*_ in the absence of an innate immune response.

Finally, we assumed an additional innate immune response, e.g., a type-I interferon response. Type-I interferons signal danger to neighboring cells, making the latter refractory to infection (*R*).(31) For simplicity, we approximated the amount of interferon in the microenvironment to be proportional to the number of infected cells.(32) Accordingly, we modeled the rate at which susceptible cells become refractory (R) by *κ*_0_*I*. Refractory cells expected to naturally revert to a susceptible state after 1/*δ*_*R*_.

#### Modeling the adaptive immune response

The model mainly focuses on the innate response. To avoid unnecessary complexity, the adaptive immune response is represented by a single term that includes two parameters: *κ*_2_, which is the maximum decay rate of infected cells due to the adaptive immune response, and τ, which represents the time post-infection that the adaptive response takes effect. This approach is similar to that in Pawelek et al.(33) This response is thus modelled using the indicator function 𝕀_*t*>τ_(t).

### Structural analysis of the model

A substantial minority of individuals with COVID-19 experience a state of sustained high inflammation or hyperinflammation. (9) We evaluated whether the model allowed the existence of such a hyperinflammatory state. We used *D*_1_ as a marker of inflammation. Through a bifurcation analysis, we searched for the existence of a stable steady state of resolved inflammation (*D*_1_=0) and the existence of a second stable steady state of hyperinflammation where *D*_1_ is maintained well above 0. This type of analysis also allows identifying basins of attraction, i.e. regions in the space of variables that lead to specific inflammation trajectories over time. We used analytical techniques combined with the numerical bifurcation software Matcont (34), a Matlab software package designed for the analysis of equations such as Eqs. 1-7.(35,36) Matcont uses continuation techniques to follow equilibria and performs normal form computations to classify bifurcation points.

### Set of virtual markers

To analyze model behavior, a set of virtual markers were computed. These markers were used to compare model predictions to clinical observations and to investigate determinants of severe COVID-19. These markers are: i) the peak viral load (peak VL, maximum value of V); ii) the time of peak viral load; iii) the difference between the viral load at its peak and the viral load 5 days after the peak; iv) peak *D*_1_ (used as a proxy measure for peak inflammation); v) the time of peak *D*_1_; vi) *D*_1_ at 60 days post-infection.

We further computed the hyperinflammation index, which takes a value of 1 if the value of log_10_(*D*_1_) at the end of the simulated period was more than 99% of its peak value, and 0 otherwise. This resulted in the categorization of each simulation into inflammation trajectory groups: i) resolved inflammation (R) or ii) hyperinflammation (H). Finally, we computed the Disease Score, defined as the total number of cells (*I*+*J*) that died over the 60-day period post-infection. This score is meant to describe disease severity, with higher scores representing more severe COVID-19.

### Identifying a space of realistic parameter values

The next step consisted in identifying a space of parameter values for which model predictions are consistent with a minimal set of clinical observations to allow *in silico* investigation of the model. In particular, acceptable parameter values should result in: i) peak viral loads (VL) values between 4 and 10 log_10_, ii) peak viral loads achieved 2 to 14 days following infection, and iii) an innate immune response during the course of infection (the activation of at least 1% of all resting innate immune cells). These are referred to as conditions i)-iii). Condition i) was chosen to represent peak viral loads observed using nasopharyngeal swabs in a population of individuals infected by the virus.(7,37) Condition ii) was chosen based on reports of peak viral loads occurring around the time of symptom onset and symptom onset primarily occurring within 14 days of infection (median 4-5 days).(14,16,32,38) Finally, we chose condition iii) to ensure *D*_1_ reaches high enough values for an observable effect of *D*_1_ on either *I*(*t*) or *J*(*t*).

The determination of the space of parameter values that satisfy the above conditions was done iteratively. First, we established bounds of values for each parameter using literature estimates when available, results from the bifurcation analysis to ensure that hyperinflammation was achievable, and preliminary simulations when no estimate was available (see Table 1). We then selected n=100,000 vectors of parameters from the parameter space using a Latin hypercube approach to minimize the chances of unexplored multidimensional subspaces.(39) Simulations were performed for each selected vector of parameters using initial conditions listed in Table 2 to predict infection and inflammation trajectories from day 0 (day of infection) to day 60. We identified regions in the parameter subspace leading to unacceptable results based on the conditions i)-iii) listed above. We subsequently refined the parameter space and repeated the procedure until we were satisfied that a randomly selected vector of parameters would lead to a high likelihood of satisfying acceptance criteria i) to iii) (>70% acceptance probability). Table 1 describes all parameters, the resulting space of parameter values along with references from the literature when applicable.

**Table 1.** Model parameters, the range of explored values used for the *in silico* investigation along with justifications and references. Par: Parameters.

| Par | Description (unit) | Range of explored values | Justification and References |
| --- | --- | --- | --- |
| $\beta$ | Infectivity rate (virion<sup>-1</sup> day<sup>-1</sup>) | [-12;-6]<br>(log<sub>10</sub> scale) | $\beta$ values of 1.9E-6 and 6.6E-7 were used in Ke et al. for the upper and lower respiratory tract, respectively.(40) A value of 5.2 x 10<sup>-6</sup> was used in Kim et al.(14) A larger range of values was used to account for variability between individuals and uncertainty in parameter values due to a possible correlation with parameter $p$. |
| $p$ | Virion production rate (virions cell<sup>-1</sup> mL<sup>-1</sup> day<sup>-1</sup>) | [-1;4]<br>(log<sub>10</sub> scale) | $p$ values of 51.4 per swab per day and 0.35 per mL per day were used in Ke et al. for the upper and lower respiratory tract, respectively.(40) A larger range of values was used to account for variability between individuals and uncertainty in parameter values due to a possible correlation with parameter $\beta$. Accordingly, it was inefficient to sample $\beta$ and $p$ independently. It was determined through linear regression that a value of log<sub>10</sub>$p$ determined by - 6.8 - 1.0*log<sub>10</sub> $\beta$ + U, where U is a uniform random variable with range [- 0.4;0.5], greatly enhanced the likelihood of peak viral loads being between 4 and 10 log<sub>10</sub> and occurring between day 2 and 14 post-infection. |
| $\delta_I$ | Death rate from viral cytopathic effects (day<sup>-1</sup>) | [0.05;0.1] | Jenner et al. used an infected cell death rate of 0.014 day<sup>-1</sup> in the preprint version of the paper.(11) The lower bound was reviewed to allow a greater probability of satisfying the simulation acceptance conditions (see Methods). A study demonstrated that viral production from infected cells was maintained at high levels for up to 6 days *in vitro*.(41) |
| $c$ | Virion clearance rate (day<sup>-1</sup>) | [10;30] | Values explored in Goncalves et al. as well as Ke et al. were between 5 and 20 day<sup>-1</sup>.(32,37) |
| $\kappa_0$ | Rate of transition to IFN induced refractory state (cell<sup>-1</sup> day<sup>-1</sup>) | [-8;-5]<br>(log<sub>10</sub> scale) | A value of 1.3 x 10<sup>-6</sup> was estimated in Ke et al.(32) |
| $\delta_R$ | Refractory state reversion rate (day<sup>-1</sup>) | [-4;-2]<br>(log<sub>10</sub> scale) | A value of 0.0044 day<sup>-1</sup> was estimated in Ke et al. (40) |
| $\tau$ | Time delay of adaptive immune response post-infection (days) | [7;40] | The range was chosen to match the variability between individuals in time of viral clearance post-infection, with a median of around 25 days.(7,40) |
| $\kappa_2$ | Effect of adaptive immune response on the clearance of infected cells (day<sup>-1</sup>) | [2;6] | This range was chosen to ensure viral clearance is achieved shortly after $\tau$ days |
| $m$ | Hill coefficient for bystander cell damage | 3 | A value of 3 ensures a steep progression of the damage rate as a function of $D_1$. In other words, we assumed that the damage due to inflammation is not substantial unless inflammation reaches high levels. |
| $\nu$ | Maximum bystander damage rate (cells day<sup>-1</sup>) | [7.5;8.5]<br>(log<sub>10</sub> scale) | An initial range of values was chosen based on bifurcation analyses. The range was refined to ensure that the around 5-20% of simulations led to hyperinflammation. |
| $K$ | $D_1$ concentration leading to half the bystander damage rate (cell) | [1.5x10<sup>6</sup>;2 x10<sup>6</sup>] | An initial range of values was chosen based on bifurcation analyses. The range was refined to ensure that the around 5-20% of simulations lead to hyperinflammation. |
| $\delta_J$ | Death rate from cell damage | [0.01;0.05] | Chosen such that the death rate for damaged cells is slower than for infected cells |
| $D_{00}$ | Resting immune cells homeostatic constant (cells mL<sup>-1</sup>) | 10<sup>6</sup> | A value of 10<sup>6</sup> was used for alveolar macrophages in Smith et al. (2011).(42) Values ranging from 4x10<sup>5</sup> and 3x10<sup>7</sup> were used for various populations of innate immune cells in Jenner et al.(11) A single value was used since we were not interested in investigating the impact of variability in $D_{00}$ on the severity of COVID-19. |
| $\lambda$ | Resting immune cells replenishing rate or recruitment rate (day<sup>-1</sup>) | [-2;1]<br>(log<sub>10</sub> scale) | A comparative value of 0.22 was used in Jenner et al. for monocytes.(11) |
| $\sigma$ | Innate immune cell activation rate (cell<sup>-1</sup> day<sup>-1</sup>) | [-8.5;-7.5]<br>(log<sub>10</sub> scale) | A value of the order of 1x10<sup>-6</sup> was used in Jenner et al.(11) The range of values was adjusted to ensure that at least 1% of all resting innate immune cells activated over the course of infection and that high inflammation levels ($D_1 > 1x10^6$) did not occur prior to peak VL. |
| $\delta_D$ | Activated innate immune cell average death rate (day<sup>-1</sup>) | [0.1;0.3] | A value of 0.3 was used for activated macrophages in Jenner et al.(11) A value of 0.04 was used for activated macrophages in Pawelek et al.(43) Natural killer cells are shown to have a turnover of around 2 weeks.(44) In Sadria et al., a value of 0.2 was used to represent the natural death rate of effector cell(13) |
| $\kappa_1$ | Effect of innate immune response on the clearance of PAMP and DAMP expressing cells (cell<sup>-1</sup> day<sup>-1</sup>) | [-8;-4]<br>(log<sub>10</sub> scale) | An initial range of values was chosen based on bifurcation analyses. The range was refined to ensure that the around 5-20% of simulations lead to hyperinflammation. |

**Table 2.** Initial conditions for all simulations.

| Variable | Description | Initial value |
| --- | --- | --- |
| $T$ | Target cells | $4.8 \times 10^8$ cells(40) |
| $R$ | Refractory cells | 0 cells |
| $I$ | Infected cells | 10 cells |
| $V$ | Virions | 0 virions |
| $J$ | Damaged cells | 0 cells |
| $D_0$ | Resting innate immune cells | $D_{00}$ (see Table 1) |
| $D_1$ | Activate innate immune cells | 0 cells |

### Simulating clinical observations

We sampled a large number of sets of parameter values (n=1,000,000) from the space of parameter values described in Table 1 using a Latin hypercube approach.(39) Accordingly, the marginal distribution of values being sampled for each parameter was uniform across the considered range (Table 1). For each selected vector, simulations were performed using initial conditions listed in Table 2 to predict infection and inflammation trajectories from day 0 (day of infection) to day 60. Analyses were only performed on the simulations satisfying the acceptance criteria (conditions i)-iii)). We first investigated the distribution of parameter values and virtual markers using histograms. We used violin plots to study bivariate associations between parameters and inflammation trajectory groups (resolved inflammation or hyperinflammation). Multivariate analyses were performed using decision trees.(45) We used the hyperinflammation index as the variable being predicted by model parameters. We first obtained a single tree. Cross-validation and deviance plots were used to guide the choice of the optimal tree.(45) To account for uncertainty around the formulation of a single tree, we also performed a Random Forest analysis, generating 100 trees, and reported the mean decrease in GINI index (a measure of decreased node impurity from choosing a parameter for tree splits, i.e. its ability to discriminate inflammation trajectory groups).(46)

### Simulating the effect of treatments on COVID-19 severity

Finally, we evaluated if the model could replicate clinical findings regarding the treatment of COVID-19. For this purpose, we selected 10,000 simulations leading to lower Disease Severity scores and the same number of simulations leading to higher scores from the sample of accepted simulations. To allow comparison with clinical data, we assumed the former group represents mild/moderate disease, while the latter represents severe COVID-19. To simulate the use of corticosteroids, we assumed a reduction of *D*_1_ by 50% for a period of 10 days. To simulate the use of potent antivirals, we decreased parameters *β* or *p* to 1% of their original value at peak viral load for the remainder of the simulation. For each simulation, we modified parameter values at the time of peak viral load as it is estimated that the peak is reached within the few days following symptom onset.(14,32,38) We also simulated treatment administration a day prior to the expected viral load peak to study the effect of early treatment. The difference in log_10_ Disease Score (with vs without treatment) was computed and reported.

## RESULTS

### The Model Allows Two Stable Steady States (Hyperinflammation and Resolved Inflammation) under Realistic Parameter Values

To understand the general dynamics of the model given by Eqs. (1) – (7) and the types of infection outcomes predicted by the model, we first performed a bifurcation analysis using baseline parameter values (see Supplementary Material for detail). Interestingly, the analysis shows that under certain biologically plausible parameter values, there exists bistability in the system with both a stable hyperinflammatory state and a stable resolved inflammatory state. In more technical terms, there are three equilibria in the model:.a stable high-inflammation state corresponding to hyperinflammation, an unstable equilibrium with non-zero, but low, inflammation and another stable equilibrium corresponding to resolved infection and resolved inflammation. The first two steady states appear/disappear following a saddle-node bifurcation as shown in the bifurcation plots (Fig S1 and S2).

The bifurcation analysis identified important parameters that dictate the existence of a hyperinflammatory steady state: i) *κ*_1_, which represents the effect of the innate response (*D*_1_) on the clearance of cells carrying PAMPs and DAMPs, and ii) the parameter *v* that dictates the amount of bystander cell damage resulting from inflammation (Fig. S1). Our analysis shows there is a threshold value of *κ*_1_ beyond which the hyperinflammatory state ceases to exist (Fig S1A). Further, hyperinflammation can only occur if there is enough immune driven inflammation beyond a threshold value of *v* (Fig S1B and Fig S2B).

When bistability exists in the system, inflammation trajectories could either converge to one stable state or the other over time. What determines the long-term inflammation trajectory is the amount of bystander cell damage *J* caused by inflammation over the course of the infection. Mathematically, this is represented by a saddle-node bifurcation. The unstable lower branch of equilibria acts as a separatrix between the hyperinflammatory and resolved-inflammation states and so defines the basin of attraction of the hyperinflammatory state (see Fig. S1). Thus, hyperinflammation can only occur if the infection induced inflammation is severe enough to force the system across this separatrix and into this basin.

Overall, these results suggest that the clearance of cells carrying PAMPs and DAMPs and the amount of bystander cell damage from inflammation are important determinants of disease outcomes. When the clearance rate of infected and damaged cells is sufficiently high or bystander cell damage is low, infection leads to non-severe outcomes.

### Model Simulations of a Virtual Cohort of Infected Individuals

We next simulated the model by sampling n=1,000,000 sets of parameter values across biologically plausible ranges (Methods). We use these simulation results to analyze the different viral load and inflammatory response trajectories in the population, such that key determinants of disease outcomes can be further identified. In our analysis, we only included simulations satisfying the acceptance criteria that are consistent with broad patterns seen in clinical studies (described in the Method section as conditions i-iii) (accepted simulations, n=739,465 of 1,000,000 or 73.9%). The distribution of parameter values that led to accepted simulations are presented in Supplementary Material (Fig. S3). Most parameters preserved their sampling distributions, i.e. the distribution remained uniform across the considered ranges after discarding simulations that do not satisfy the acceptance criteria. Further, there was little correlation between almost all parameters. However, there was a strong correlation between *β* and *p* (−0.99). Consequently, parameters *β* and *p* could not be independently sampled in order to output acceptable simulations (Fig. S4). Table 1 describes the sampling strategy used to account for this dependency.

### A Strong Association Between Hyperinflammation & the Disease Score

To represent disease severity, we define the Disease score as the total number of infected and bystander cells *I* and *J*, that died over a 60-day period. Simulations give rise to the Disease Score distribution are illustrated in Fig. 2. They are categorized in groups by inflammation trajectory: i) the inflammation marker *D*_1_ decreased after peak inflammation or ii) the inflammation marker *D*_1_ kept increasing or was maintained at high level following peak inflammation (hyperinflammation). There was a direct link between the presence of hyperinflammation and the Disease Score (see Fig. 2A). Overall, 13.5% of accepted simulations exhibited hyperinflammation. Hyperinflammation and a high Disease Score were both associated with a higher number of cells that died following injury from inflammation (Fig. 2B).

**Figure 2.**
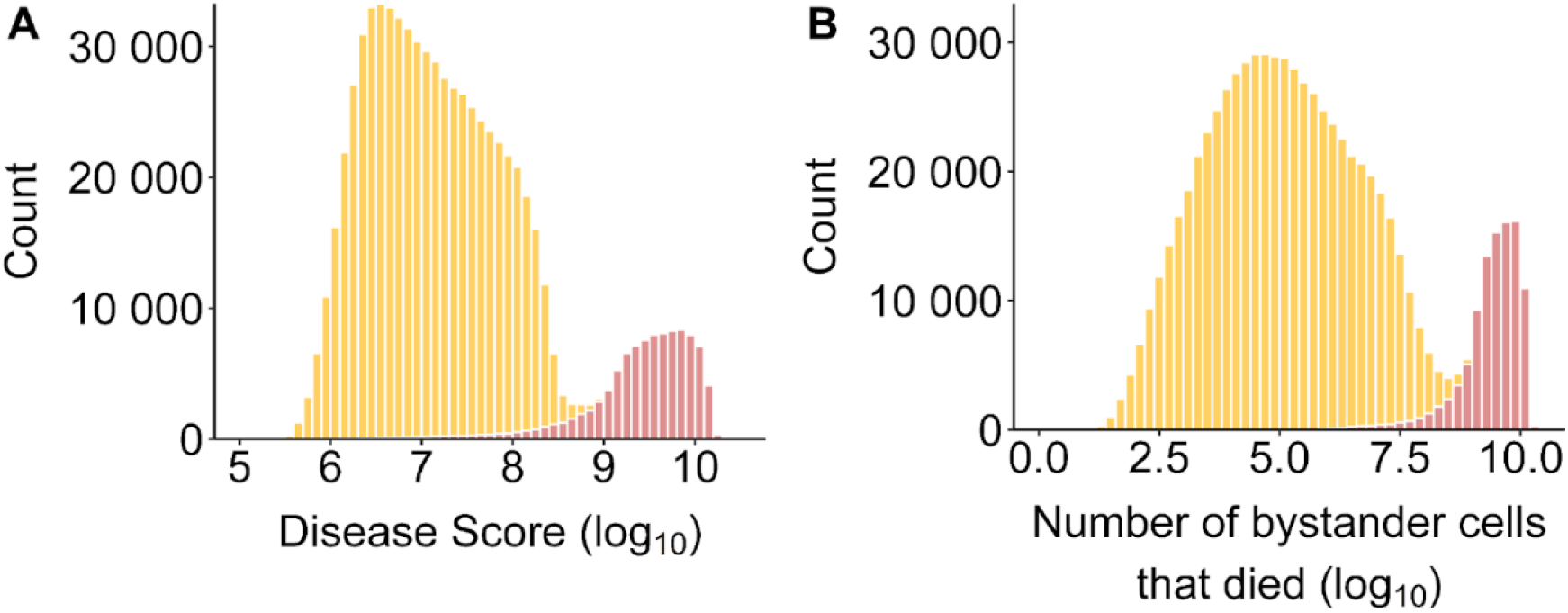
(A) Distribution of Disease Scores and (B) distribution of the total number of bystander cells that died by inflammation trajectory groups: Resolved inflammation (orange), Hyperinflammation (pink).

### Similar Viral Load but Divergent Inflammation Trajectories

Viral load and inflammation trajectories over time by inflammation groups are presented in Fig. 3. Simulations resulting in both resolved inflammation or hyperinflammation exhibited similar viral load dynamics. The distribution of peak viral load and the time to reach this peak after infection largely overlapped between groups. However, those with resolved inflammation tended to have a faster VL decay after peak VL (Fig. 3C). The most remarkable difference between groups pertained to the dynamics of the inflammation marker *D*_1_. Peak levels of inflammation were higher for simulations resulting in hyperinflammation. For those with resolved inflammation, peak inflammation was generally observed around the time of peak viral load while for those with hyperinflammation, peak inflammation was observed much later (Fig. 3H).

**Figure 3.**
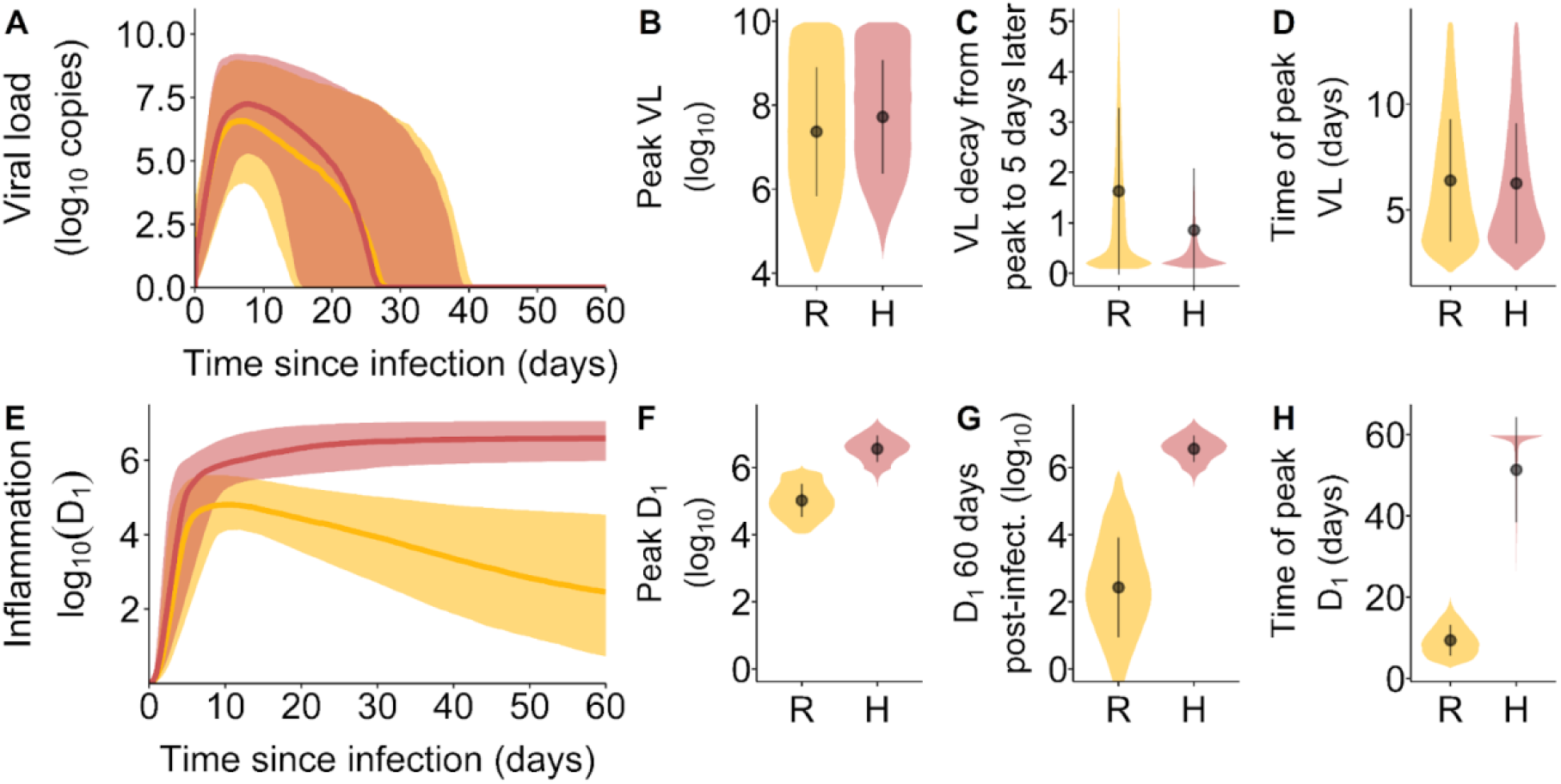
Viral load and inflammation trajectory characteristics by inflammation trajectory groups. (A) Viral loads over the course of infection. The shaded area corresponds to the 10th and 90th percentiles of the viral loads, while the curve represents the median. (B) The distribution of peak viral loads, (C) the VL decay from peak infection to 5 days after peak infection and (D) the time of occurrence of peak VL after infection. (E) Inflammation trajectories by inflammation trajectory groups. The shaded area corresponds to the 10th and 90th percentiles of *D*_1_, while the curve represents the median. (F) the distribution of peak *D*_1_, (G) the distribution of *D*_1_ at 60 days post-infection and (H) the time of occurrence of peak *D*_1_ after infection. In orange and represented by the symbol R, Resolved inflammation. In pink and represented by the symbol H, Hyperinflammation.

### Association between Hyperinflammation and Characteristics of the Innate Immune Response

Next, we compared the distribution of parameter values between inflammation trajectory groups. Figure 4 shows violin plots for the 15 parameters that were allowed to vary between simulations. The most striking differences between groups are observed for parameters *κ*_1_, *λ, κ*_0_, *σ, δ*_*D*_ and *v*. Lower values of *κ*_0_ (type-I IFN secretion and/or response) and *κ*_1_ (cytolytic and phagocytic activities of innate immune cells) were associated with a greater risk of hyperinflammation and a high Disease Score. Similarly, higher values of *λ* (innate immune cell recruitment rate), *σ* (activation rate), 1/*δ*_*D*_ (survivability of activated innate immune cells) and *v* (damage rate due to inflammation) were associated with hyperinflammation and a high Disease Score.

**Figure 4.**
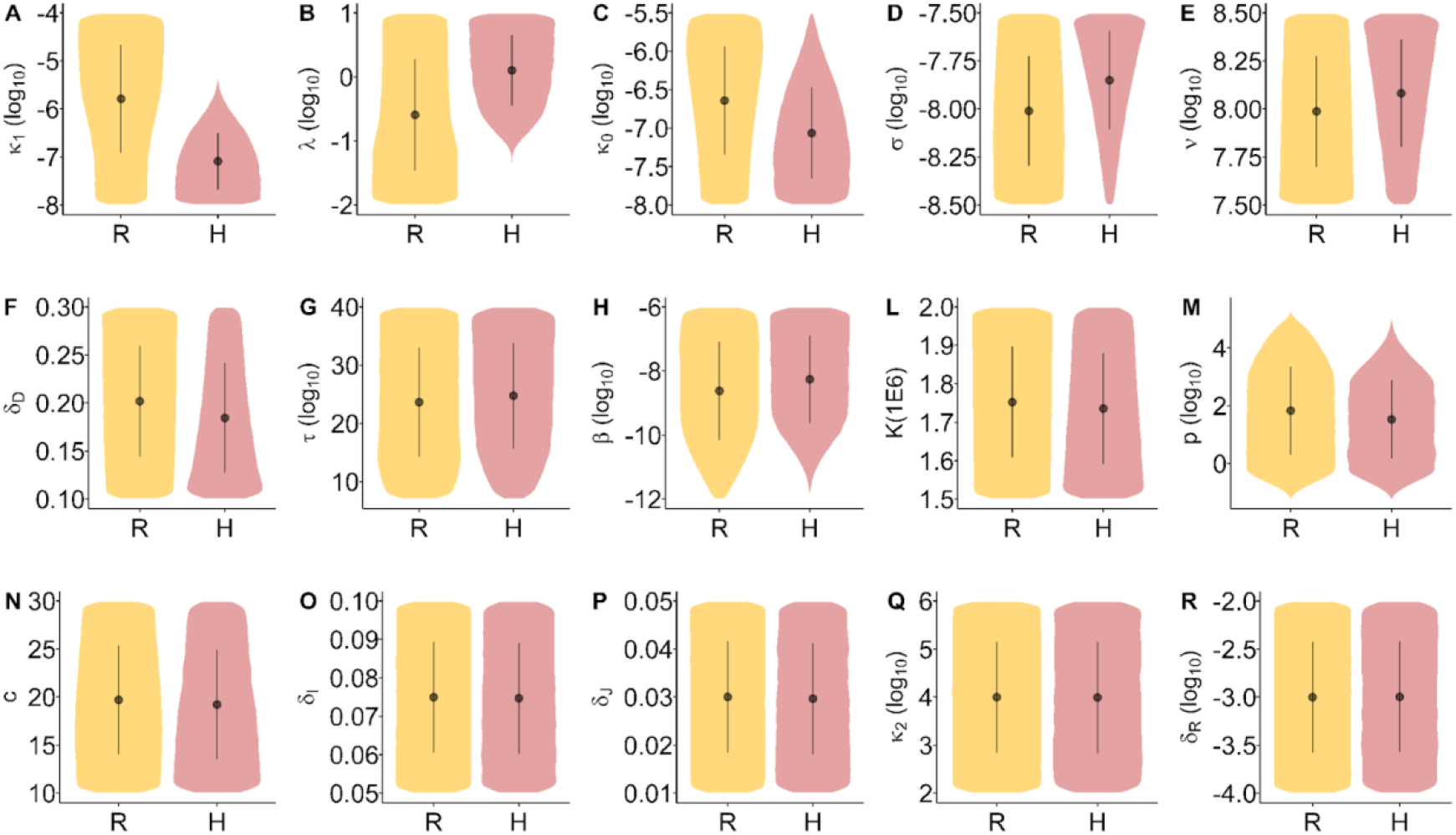
Distribution of the model parameters by inflammation trajectory groups. Distribution overlap may be discriminated from multivariate models. Inflammation trajectory groups R: Resolved inflammation, H: Hyperinflammation.

### Prediction of the Risk of Hyperinflammation from Characteristics of the Innate Immune Response

We used regression tree analysis to reveal the discriminatory importance of parameters from a multivariate perspective. The regression trees attempted to discriminate simulations leading to hyperinflammation from those leading to a resolved inflammatory state. Results are shown in Fig. 5.

**Figure 5.**
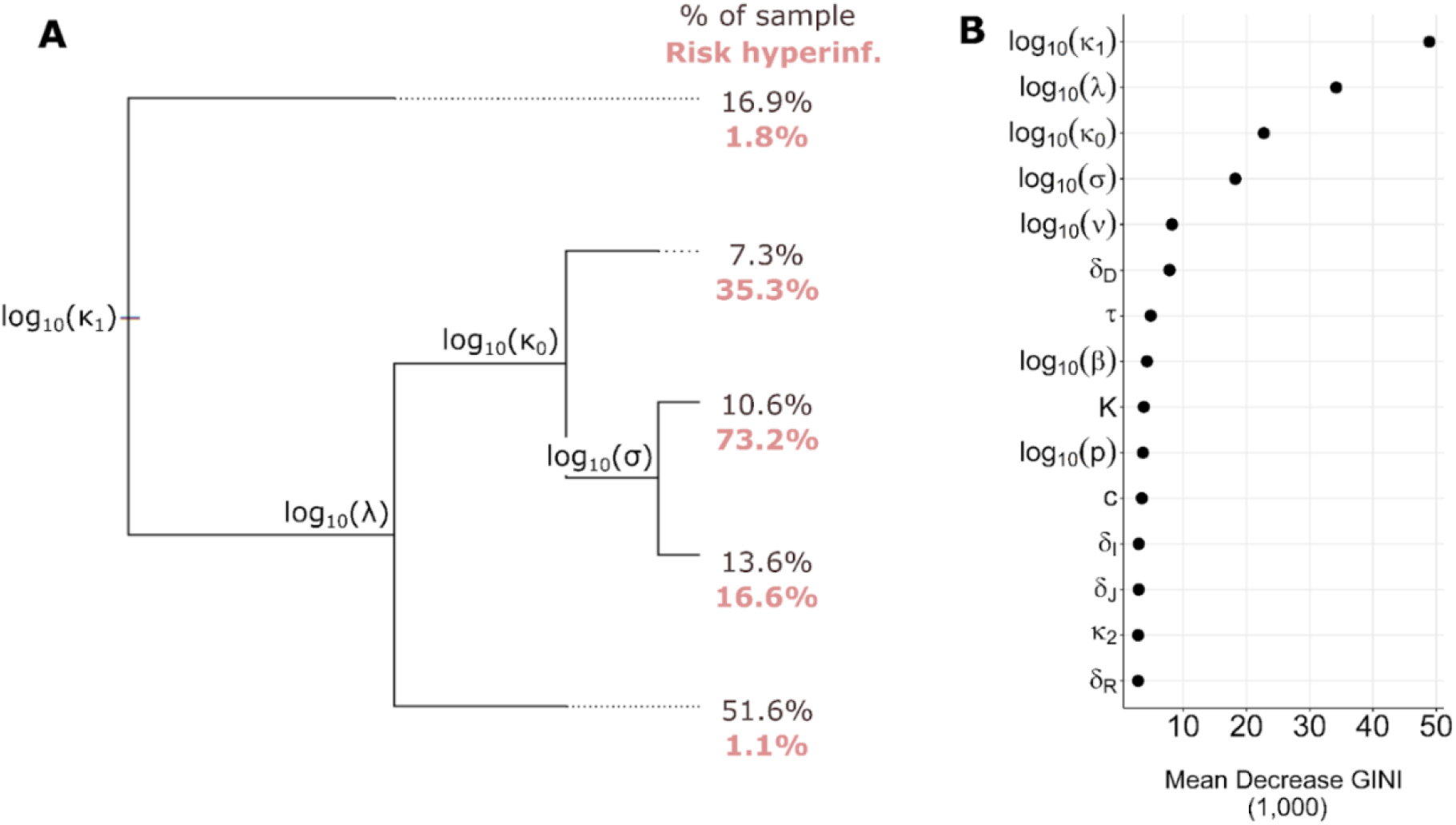
Regression tree analysis results. (A) Single optimal tree for the prediction of hyperinflammation from model parameters. The tree reads from left to right. At each labeled node, simulations either go up if the value for the associated parameter is higher than a threshold determined by the procedure (threshold not shown, see Supplementary Figure S7), or down otherwise. Branch length represents the amount of classification error explained by the node. At each terminal node, the percentage of simulations as well as the risk of hyperinflammation within members of the node are reported. (B): Parameter importance based on the GINI index. A greater mean GINI decrease indicates a parameter that is more discriminatory.

The tree has a root (left side), branches, nodes (where branches separate) and terminal leaves (right side). Simulations enter at the root and separate at nodes. At each node, one parameter and one threshold value were chosen by the regression algorithm based on their ability to separate simulations into more homogeneous groups in terms of inflammation trajectory. Simulations are directed toward the lower branch if the value for the chosen parameter for the simulation is smaller than the threshold, or toward the upper branch otherwise. This resulted in 5 terminal leaves that partitioned all simulations into 5 groups. Longer branches following a node signify that the node allowed a better separation of the two inflammation trajectory groups.

Fig. 5 illustrates the multivariate conditions favoring hyperinflammation. The most important parameters pertain to type-I IFN response, the ability of the system to clear cells carrying PAMPs and DAMPs (*κ*_0_ and *κ*_1_) and the immune cell activation and recruitment rate (*σ* and *λ*). In particular, combined low values of *κ*_0_ and *κ*_1_ and high values of *σ* and *λ* led to a dramatically increased chance of hyperinflammation (73.2% risk of hyperinflammation).

### Differences in the Effect of Treatments in terms of Treatment Type, Administration Time and Predicted Disease Severity

To validate and demonstrate the utility of our model, we simulated the use of corticosteroids and antivirals in infected individuals and compared the model results with clinical findings. First, we modeled corticosteroid treatment by assuming the treatment leads to a reduction of *D*_1_ by 50% for a period of 10 days following peak infection. The *in silico* administration of corticosteroids had a remarkably different effect on the Disease Score depending on the inflammation trajectory groups (Fig. 6). Among those simulations where resolved inflammation is predicted in the absence of treatment (orange plot), corticosteroids were often detrimental (32% chance of an increase in Disease Score). Such a detrimental effect was not observed among those for which hyperinflammation was predicted in absence of treatment (pink plot). Greater improvements were observed in the latter group: 23% had a greater than 0.5 log_10_ decrease in Disease Score, compared to only 0.7% in the former group. There were only small differences in the effect of corticosteroids across the investigated timing of drug administration (at peak viral load in Fig 6A and a day prior in Fig 6B). Corticosteroid treatment also had an effect on the slope of viral load decay, which was substantial for those simulations of mild/moderate disease and lesser among simulations of severe COVID-19 (Fig S8).

**Figure 6.**
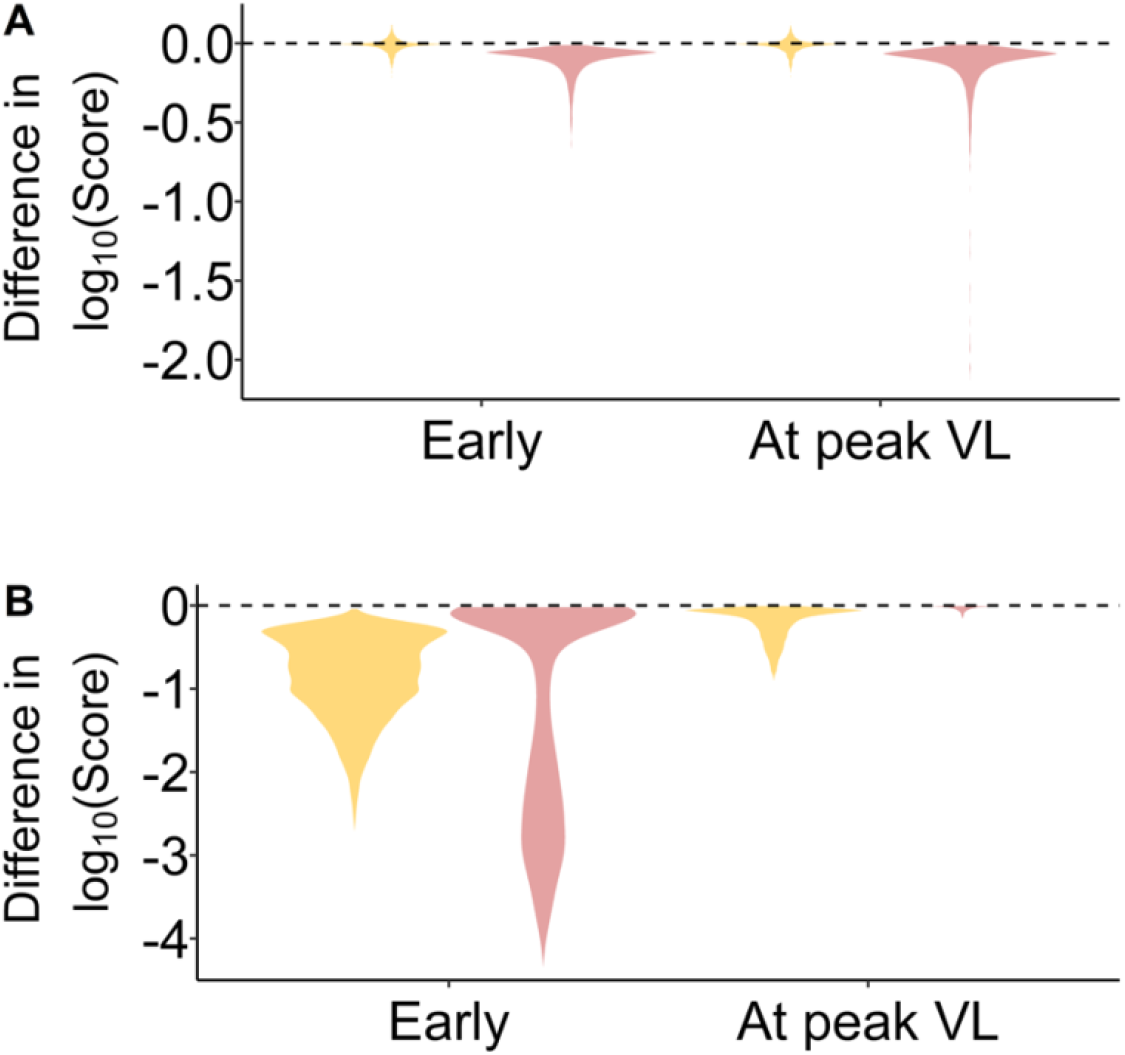
Violin plots of the effect of virtual treatment on the Disease Score. A) represents administration of corticosteroids while B) represents antiviral drug administration. Negative values represent improvements while positive values represent the worsening of symptoms. Note there were no clear difference between a reduction in *β* or *p* in the simulation of antivirals so the figure applies to both cases. Orange denotes the effect of treatment among individuals who would have resolved inflammation in the absence of treatment, whereas pink denotes the effect of treatment among individuals who would have had hyperinflammation in the absence of treatment.

Simulations of the effect of antivirals were performed by decreasing parameters *p* or *β* to 1% of their original value at peak viral load. Modifying *p* or *β* had a very similar effect on

Disease Scores. None of the simulations revealed a substantial increase in Disease Score. However, the impact of antivirals was very different between inflammation trajectory groups and between times of drug administration. Earlier drug administration led to large improvements in Disease Score for both groups (Fig 6B). However, administration at peak viral load led to much reduced improvements in Disease Score (Fig 6A), in particular for those simulations leading to hyperinflammation in absence of treatment. In this case, infection had already driven inflammation to high levels by the time peak viral load is reached (although peak inflammation was not reached until much later), which translated into a number of damaged cells that corresponded to being within the basin of attraction of hyperinflammation (Fig S1).

## DISCUSSION

In this work, a mathematical model was formulated to represent the within-host dynamics of COVID-19 infection and inflammation. The objective was to provide a quantitative explanation for the range of COVID-19 symptom severity among individuals and to reveal the discriminatory importance of modeled mechanisms. The hypothesis we explored was that high levels of inflammation in COVID-19 may produce a significant amount of damage to uninfected cells. These cells would then produce DAMPs that further stimulate the inflammatory response. This model produced predictions that are consistent with clinical observations.

### Hyperinflammation and disease severity

The model predicts that those with higher Disease Scores had substantially higher levels of inflammation (Fig 2A). Further, peak inflammation was not reached until much later in those simulations leading to hyperinflammation (Fig 3H). These modeling results are consistent with clinical findings that revealed non-survivors tend to experience hyperinflammation and increasing levels of inflammatory biomarkers up to the time of death.(7,9,47) Our analysis provides an explanation of these findings. In particular, the numerical bifurcation analysis suggests that viral infection can push the immune system into a self-sustaining high inflammatory state that can persist well beyond the resolution of infection. This hyperinflammatory state causes additional damage to uninfected cells, consequently leading to much higher Disease Scores. Due to the strong association between hyperinflammation and the Disease Score, we used those simulations that led to hyperinflammation as an *in silico* description of severe COVID-19. The percentage of simulations exhibiting hyperinflammation (13.5%) closely matched the reported proportion of the infected population with severe COVID-19 (14%), further motivating this decision.(1)

### Viral load dynamics and disease severity

In terms of viral load, the main difference that was reported between severe COVID-19 and those with milder disease pertained to the slope of VL after disease onset.(7) In particular, a faster VL decay was observed for those with milder disease after symptom onset, as measured using nasopharyngeal swabs.(7) This finding was also reported for VL sampled using other means and from various physiological compartments.(48) Interestingly, disease severity does not correlate strongly with peak viral loads.(7,48) Our simulation results are consistent with these findings (Fig. 3B and 3C).The model offers an explanation to the slower viral load decay after peak infection in cases of severe COVID-19 through parameter *κ*_1_; a lower *κ*_1_ value both results in slower clearance of productively infected cells (see Eq. 3) and increases the risk of hyperinflammation (see Fig 5). In fact, having a low *κ*_1_ value was the most important predictor of hyperinflammation (Fig 5B). In the model, low *κ*_1_ values lead to the persistence of *I* and *J* cells, thereby prolonging PAMP and DAMP signaling and its downstream impact on the inflammatory response. The heightened inflammatory response promotes the generation of damaged cells *J*, strengthening DAMP signaling. Our analysis suggests that the ensuing feedback loop is the hallmark of hyperinflammation and severe COVID-19.

### Characteristics of the innate immune response and disease severity

Many clinically observed associations were found between components of the innate immune response and disease severity. Among them, a dysregulated IFN response has been repeatedly associated with severe COVID-19.(10,49) Comparatively, our model suggests that a weaker IFN response (lower *κ*_0_) leads to a higher likelihood of hyperinflammation (see Fig 4C, 5A and 5B). One of the roles of type-I IFN is to limit the number of target cells that can be infected.(31) Although it did not have a big impact on peak VL or the time to reach this peak in our simulations, an efficient type-I IFN response (higher *κ*_0_ values) had a significant impact on inflammation. IFN by limiting the rate and shear number of cells that carry PAMPs also constrains inflammation.

Poor NK-cell cytotoxic ability was also linked with severe COVID-19.(50) NK-cells are important actors in the innate immune response that can target both infected and damaged cells and release molecules that precipitate their apoptosis.(29,30) In the model, poorer clearance of cells having PAMPs and DAMPs by the innate immune response is represented by lower *κ*_1_ values, the most important predictor of hyperinflammation and severe COVID-19 (Fig 5B). A poorer response from NK cells could hence lead to severe COVID-19 by enabling prolonged PAMP and DAMP signaling. Patients with severe COVID-19 also have increased levels of neutrophil extracellular traps (NETs).(25,26) These can lead to immunothrombosis and the generation of damaged cells.(23,24) In the model, higher generation of damaged cells is represented by larger values of the parameter *v* (Eq 5). The bifurcation analysis suggests that larger values of *v* lowers the threshold for the number of damaged cells that are required to reach a hyperinflammatory state (Fig S1), facilitating severe COVID-19.

Finally, clinically defined severe COVID-19 has been associated with higher abundance and activation of proinflammatory macrophages.(51) In the model, those with higher disease scores had a higher number of innate immune cells (*D*_0_ + *D*_1_) and a higher proportion of these cells were activated. The parameters that dictate innate immune cell recruitment (*λ*) and activation (*σ*) were both strongly associated with hyperinflammation and having a high Disease Score.

### Effect of corticosteroids and antivirals on disease severity

We evaluated if the model could replicate clinical findings regarding the treatment of COVID-19. We used the model to simulate the treatment of corticosteroid, such as dexamethasone, or an effective antiviral. The model predictions are consistent with many clinical findings, suggesting that the model we developed here will be a useful tool to understand SARS-CoV-2 pathogenesis and predicting the impact of treatment. Indeed, observational studies revealed the use of corticosteroids can lead to more severe symptoms in those with milder disease, but generally improves outcomes for those with more severe symptoms.(51,53–56) Further, it is reported that those with milder COVID-19 generally experience slower viral load decay under corticosteroid treatment, an effect that was not found to be statistically significant among those with severe disease.(53) This latter result was also observed in our simulations (Fig S8). One of the roles of inflammation is to stimulate cytolytic and phagocytic activities. By lowering this ability among those who experienced milder disease, the use of corticosteroids may lead to slower viral clearance. Hence, corticosteroids could have both a negative effect (slower clearance of PAMP carrying cells) and a beneficial effect (slower rate of bystander cell damage) on disease pathogenesis in those with mild disease. This could result in corticosteroids sometimes improving, sometimes worsening disease severity. Comparatively, the net beneficial effect of corticosteroids on those with severe COVID-19 may be the result of a smaller downstream impact of the drug on already less efficient NK cells.(50)

For antivirals, simulations suggest that early administration is crucial for antivirals to impact disease severity, particularly for those that would have experienced severe COVID-19 in absence of treatment (Fig 6). Our results also suggest the existence of an inflammation threshold beyond which antivirals may be unable to prevent hyperinflammation. It also suggests that early administration could reduce disease severity by preventing the infection from driving inflammation across the threshold. Comparatively, many clinical trials failed to show a substantial effect of antivirals.(57) More recently, a reduction of around 50% in the risk of hospitalization was observed after administering the antiviral molnupiravir.(58) The studied cohort consisted of individuals that had mild/moderate symptoms and were not expected to be hospitalized within 48 hours of randomization.(58) Our results suggest this latter criteria may be crucial to ensure the beneficial effect of antivirals on disease severity.

### Other within-host models of SARS-CoV-2 infections

The model we propose is unique, but many of the effects it includes are present in other within-host models of SARS-CoV-2 infection. Despite differences in model formulations, there is a general agreement across models about the necessity for early antiviral administration.(12–15) However, models differed in terms of the components of the inflammatory, innate or adaptive immune response they include.(11–17) Some of the models included an effect of type-I interferon, either as a promoter of the pro-inflammatory response or helping susceptible cells resist infection.(11–13) One of these models concluded, as per our analysis, that an inefficient type-I IFN response can lead to accentuated tissue damage.(11) In comparison, our model simultaneously explains the more important clinical associations between severe disease and biomarkers of the innate immune response. It distinguishes itself by the inclusion of both a positive feedback loop between damaged cells and inflammation and an effect of innate immune cells on both infected and damaged cells. This latter effect highlights the importance of cells capable of clearing both types of cells in the pathogenesis of severe COVID-19. Although we investigated more complex models, we decided against the modeling of individual cytokines or cells, as they often exhibit overlapping functions, and because some of these functions have been poorly studied leading to uncertainties in parameter values. However, some of the more complex models reported in the literature did give rise to interesting hypotheses that may warrant further investigation, such as the role of monocyte-to-macrophage differentiation or the role of anti-inflammatory cytokines.(11,12)

## Conclusion

Our analysis revealed key aspects of the innate immune response that dictate inflammation trajectories and disease severity. The most important parameter suggested by bifurcation and decision tree analyses was *κ*_1_, representing the ability of the system to rid itself of cells carrying PAMPs and DAMPs. The analysis suggested that when this parameter is high enough, hyperinflammation can be avoided. The bifurcation analysis also suggested that small values of *v*, representing the amount of bystander cell damage due to inflammation has a similar effect. In other words, the ability of the innate immune response to target PAMPs and DAMPs carrying cells appears to be key to the determination of COVID-19 severity. This suggests that therapies that specifically target aspects of the innate immune response may prove beneficial in comparison to broadly acting anti-inflammatory agents. The model also underlined the role of DAMPs in maintaining high levels of inflammation later in the course of infection. It suggests DAMPs may be an interesting therapeutic target for COVID-19. When exploring such novel treatment strategies, the model presented here could provide a means of exploring timing of treatment and dose effects *in silico*. Hopefully, a better understanding of the pathology of SARS-CoV-2 will lead to decreased mortality in this and similar diseases.

## Data Availability

All data produced in the present work are contained in the manuscript.

## Acknowledgements

This work was performed under the auspices of the US Department of Energy through Los Alamos National Laboratory, which is operated by Triad National Security, LLC for the National Nuclear Security Administration of the U.S. Department of Energy (contract No. 89233218CNA000001). The work was supported by the Laboratory Directed Research and Development program of Los Alamos National Laboratory (projects No. 20200743ER, 20200695ER, and 20210730ER), by NIH grants R01-AI028433, R01-OD011095 (ASP), R01-AI15270301 (RK), by the Defense Advanced Research Projects Agency (contract No. W911NF-17-2-0034) and by the DOE Office of Science through the National Virtual Biotechnology Laboratory, a consortium of DOE National Laboratories focused on response to COVID-19, with funding provided by the Coronavirus CARES Act. Additional support was provided by the Center for Nonlinear Studies at Los Alamos National Laboratory.

## Competing interests

The authors have declared that no competing interests exist.

## Notes

### Competing Interest Statement

The authors have declared no competing interest.

## REFERENCES

1. Gandhi RT, Lynch JB, Del Rio C. Mild or moderate Covid-19. New England Journal of Medicine. 2020;383(18):1757–66.

2. Avila J, Long B, Holladay D, Gottlieb M. Thrombotic complications of COVID-19. The American Journal of Emergency Medicine. 2020;39:213–8.

3. Gao Y, Ding M, Dong X, Zhang J, Kursat Azkur A, Azkur D, et al. Risk factors for severe and critically ill COVID-19 patients: a review. Allergy. 2021;76(2):428–55.

4. Fox SE, Akmatbekov A, Harbert JL, Li G, Brown JQ, Vander Heide RS. Pulmonary and cardiac pathology in African American patients with COVID-19: an autopsy series from New Orleans. The Lancet Respiratory Medicine. 2020;8(7):681–6.

5. Bryce C, Grimes Z, Pujadas E, Ahuja S, Beasley MB, Albrecht R, et al. Pathophysiology of SARS-CoV-2: the Mount Sinai COVID-19 autopsy experience. Modern Pathology. 2021;34(8):1456–67.

6. Del Valle DM, Kim-Schulze S, Huang H-H, Beckmann ND, Nirenberg S, Wang B, et al. An inflammatory cytokine signature predicts COVID-19 severity and survival. Nature Medicine. 2020;26(10):1636–43.

7. Lucas C, Wong P, Klein J, Castro TB, Silva J, Sundaram M, et al. Longitudinal analyses reveal immunological misfiring in severe COVID-19. Nature. 2020;584(7821):463–9.

8. Liao J, Wen B, Deng X. Progress on role of cytokine storm in exacerbation of coronavirus disease 2019 (COVID-19): Review. Chinese Journal of Cellular and Molecular Immunology. 2020;36(10):941–7.

9. Gustine JN, Jones D. Immunopathology of Hyperinflammation in COVID-19. The American Journal of Pathology. 2020;191(1):4–17.

10. Blanco-Melo D, Nilsson-Payant BE, Liu W-C, Uhl S, Hoagland D, Møller R, et al. Imbalanced host response to SARS-CoV-2 drives development of COVID-19. Cell. 2020;181(5):1036-1045. e9.

11. Jenner AL, Aogo RA, Alfonso S, Crowe V, Deng X, Smith AP, et al. COVID-19 virtual patient cohort suggests immune mechanisms driving disease outcomes. PLoS pathogens. 2021;17(7):e1009753.

12. Mochan E, Sego TJ, Gaona L, Rial E, Ermentrout GB. Compartmental Model Suggests Importance of Innate Immune Response to COVID-19 Infection in Rhesus Macaques. Bulletin of Mathematical Biology. 2021;83(7):1–26.

13. Sadria M, Layton AT. Modeling Within-host SARS-CoV-2 Infection Dynamics and Potential Treatments. Viruses. 2021;13(6):1141.

14. Kim KS, Ejima K, Iwanami S, Fujita Y, Ohashi H, Koizumi Y, et al. A quantitative model used to compare within-host SARS-CoV-2, MERS-CoV, and SARS-CoV dynamics provides insights into the pathogenesis and treatment of SARS-CoV-2. PLoS biology. 2021;19(3):e3001128.

15. Cao Y, Gao W, Caro L, Stone JA. Immune-Viral Dynamics Modeling for SARS-CoV-2 Drug Development. Clinical and Translational Science. 2021;

16. Néant N, Lingas G, Le Hingrat Q, Ghosn J, Engelmann I, Lepiller Q, et al. Modeling SARS-CoV-2 viral kinetics and association with mortality in hospitalized patients from the French COVID cohort. Proceedings of the National Academy of Sciences. 2021;118(8).

17. Goyal A, Cardozo-Ojeda EF, Schiffer JT. Potency and timing of antiviral therapy as determinants of duration of SARS-CoV-2 shedding and intensity of inflammatory response. Science Advances. 2020;6(47):eabc7112.

18. Carty M, Guy C, Bowie AG. Detection of viral infections by innate immunity. Biochemical Pharmacology. 2020;183:114316.

19. Zindel J, Kubes P. DAMPs, PAMPs, and LAMPs in immunity and sterile inflammation. Annual Review of Pathology: Mechanisms of Disease. 2020;15:493–518.

20. Poljšak B, Milisav I. Clinical implications of cellular stress responses. Bosnian Journal of Basic Medical Sciences. 2012;12(2):122.

21. Shen H, Kreisel D, Goldstein DR. Processes of sterile inflammation. The Journal of Immunology. 2013;191(6):2857–63.

22. Murao A, Aziz M, Wang H, Brenner M, Wang P. Release mechanisms of major DAMPs. Apoptosis. 2021;26(3):152–62.

23. Frantzeskaki F, Armaganidis A, Orfanos SE. Immunothrombosis in acute respiratory distress syndrome: cross talks between inflammation and coagulation. Respiration. 2017;93(3):212–25.

24. Eltzschig HK, Carmeliet P. Hypoxia and inflammation. New England Journal of Medicine. 2011;364(7):656–65.

25. Middleton EA, He X-Y, Denorme F, Campbell RA, Ng D, Salvatore SP, et al. Neutrophil extracellular traps contribute to immunothrombosis in COVID-19 acute respiratory distress syndrome. Blood. 2020;136(10):1169–79.

26. Zuo Y, Yalavarthi S, Shi H, Gockman K, Zuo M, Madison JA, et al. Neutrophil extracellular traps in COVID-19. JCI Insight. 2020;5(11).

27. Porto BN, Stein RT. Neutrophil extracellular traps in pulmonary diseases: too much of a good thing? Frontiers in Immunology. 2016;7:311.

28. Adib-Conquy M, Scott-Algara D, Cavaillon J-M, Souza-Fonseca-Guimaraes F. TLR-mediated activation of NK cells and their role in bacterial/viral immune responses in mammals. Immunology and Cell Biology. 2014;92(3):256–62.

29. Topham NJ, Hewitt EW. Natural killer cell cytotoxicity: how do they pull the trigger? Immunology. 2009;128(1):7–15.

30. Borchers MT, Harris NL, Wesselkamper SC, Vitucci M, Cosman D. NKG2D ligands are expressed on stressed human airway epithelial cells. American Journal of Physiology-Lung Cellular and Molecular Physiology. 2006;291(2):L222–31.

31. Sallard E, Lescure F-X, Yazdanpanah Y, Mentre F, Peiffer-Smadja N. Type 1 interferons as a potential treatment against COVID-19. Antiviral Research. 2020;178:104791.

32. Ke R, Zitzmann C, Ho DD, Ribeiro R, Perelson AS. In vivo kinetics of SARS-CoV-2 infection and its relationship with a person’s infectiousness. MedRxiv. 2021;

33. Pawelek KA, Huynh GT, Quinlivan M, Cullinane A, Rong L, Perelson AS. Modeling within-host dynamics of influenza virus infection including immune responses. PLoS Computational Biology. 2012;8(6):e1002588.

34. Dhooge A, Govaerts W, Kuznetsov YA, Meijer HGE, Sautois B. New features of the software MatCont for bifurcation analysis of dynamical systems. Mathematical and Computer Modelling of Dynamical Systems. 2008;14(2):147–75.

35. Breda D, Diekmann O, Liessi D, Scarabel F. Numerical bifurcation analysis of a class of nonlinear renewal equations. Electronic Journal of Qualitative Theory of Differential Equations. 2016;65:1–24.

36. De Souza DC, Craig M, Cassidy T, Li J, Nekka F, Bélair J, et al. Transit and lifespan in neutrophil production: implications for drug intervention. Journal of Pharmacokinetics and Pharmacodynamics. 2018;45(1):59–77.

37. Gonçalves A, Bertrand J, Ke R, Comets E, De Lamballerie X, Malvy D, et al. Timing of antiviral treatment initiation is critical to reduce SARS-CoV-2 viral load. CPT: Pharmacometrics & Systems Pharmacology. 2020;9(9):509–14.

38. Cevik M, Tate M, Lloyd O, Maraolo AE, Schafers J, Ho A. SARS-CoV-2, SARS-CoV, and MERS-CoV viral load dynamics, duration of viral shedding, and infectiousness: a systematic review and meta-analysis. The Lancet Microbe. 2020;2(1):E13–22.

39. Huntington DE, Lyrintzis CS. Improvements to and limitations of Latin hypercube sampling. Probabilistic Engineering Mechanics. 1998;13(4):245–53.

40. Ke R, Zitzmann C, Ribeiro RM, Perelson AS. Kinetics of SARS-CoV-2 infection in the human upper and lower respiratory tracts and their relationship with infectiousness. MedRxiv. 2020;

41. Zhu N, Wang W, Liu Z, Liang C, Wang W, Ye F, et al. Morphogenesis and cytopathic effect of SARS-CoV-2 infection in human airway epithelial cells. Nature Communications. 2020;11(1):1–8.

42. Smith AM, McCullers JA, Adler FR. Mathematical model of a three-stage innate immune response to a pneumococcal lung infection. Journal of Theoretical Biology. 2011;276(1):106–16.

43. Pawelek KA, Dor Jr D, Salmeron C, Handel A. Within-host models of high and low pathogenic influenza virus infections: The role of macrophages. PloS One. 2016;11(2):e0150568.

44. Hervier B, Russick J, Cremer I, Vieillard V. NK cells in the human lungs. Frontiers in Immunology. 2019;10:1263.

45. Breiman L, Friedman JH, Olshen RA, Stone CJ. Classification and regression trees. Routledge; 2017.

46. Cutler A, Cutler DR, Stevens JR. Random forests. In: Ensemble Machine Learning. Springer; 2012. p. 157–75.

47. Zhou F, Yu T, Du R, Fan G, Liu Y, Liu Z, et al. Clinical course and risk factors for mortality of adult inpatients with COVID-19 in Wuhan, China: a retrospective cohort study. The Lancet. 2020;395(10229):1054–62.

48. Wang Y, Zhang L, Sang L, Ye F, Ruan S, Zhong B, et al. Kinetics of viral load and antibody response in relation to COVID-19 severity. The Journal of Clinical Investigation. 2020;130(10).

49. Hadjadj J, Yatim N, Barnabei L, Corneau A, Boussier J, Smith N, et al. Impaired type I interferon activity and inflammatory responses in severe COVID-19 patients. Science. 2020;369(6504):718–24.

50. Zheng M, Gao Y, Wang G, Song G, Liu S, Sun D, et al. Functional exhaustion of antiviral lymphocytes in COVID-19 patients. Cellular & Molecular Immunology. 2020;17(5):533–5.

51. Liao M, Liu Y, Yuan J, Wen Y, Xu G, Zhao J, et al. Single-cell landscape of bronchoalveolar immune cells in patients with COVID-19. Nature Medicine. 2020;26(6):842–4.

52. Beigel JH, Tomashek KM, Dodd LE, Mehta AK, Zingman BS, Kalil AC, et al. Remdesivir for the treatment of Covid-19—preliminary report. New England Journal of Medicine. 2020;173(2).

53. Li Q, Li W, Jin Y, Xu W, Huang C, Li L, et al. Efficacy evaluation of early, low-dose, short-term corticosteroids in adults hospitalized with non-severe COVID-19 pneumonia: a retrospective cohort study. Infectious Diseases and Therapy. 2020;9(4):823–36.

54. Horby P, Mafham M, Linsell L, Bell JL, Staplin N, Emberson JR, et al. Effect of hydroxychloroquine in hospitalized patients with COVID-19: preliminary results from a multi-centre, randomized, controlled trial. MedRxiv. 2020;

55. Keller MJ, Kitsis EA, Arora S, Chen J-T, Agarwal S, Ross MJ, et al. Effect of systemic glucocorticoids on mortality or mechanical ventilation in patients with COVID-19. Journal of Hospital Medicine. 2020;15(8):489–93.

56. Zha L, Li S, Pan L, Tefsen B, Li Y, French N, et al. Corticosteroid treatment of patients with coronavirus disease 2019 (COVID-19). Medical Journal of Australia. 2020;212(9):416–20.

57. Tarighi P, Eftekhari S, Chizari M, Sabernavaei M, Jafari D, Mirzabeigi P. A review of potential suggested drugs for coronavirus disease (COVID-19) treatment. European Journal of Pharmacology. 2021;895:173890.

58. Merck. Merck and Ridgeback’s Investigational Oral Antiviral Molnupiravir Reduced the Risk of Hospitalization or Death by Approximately 50 Percent Compared to Placebo for Patients with Mild or Moderate COVID-19 in Positive Interim Analysis of Phase 3 Study. [cited 2021 Oct 19]; Available from: https://www.merck.com/news/merck-and-ridgebacks-investigational-oral-antiviral-molnupiravir-reduced-the-risk-of-hospitalization-or-death-by-approximately-50-percent-compared-to-placebo-for-patients-with-mild-or-moderat/

