## Supplementary Material for "A simple model of COVID-19 explains disease severity and the effect of treatments"

### Mathematical analyses of the model

Here, we detail the analytical results discussed in the main text with the goal of understanding the long-term behavior of the within host model described in the main text

$$\left.\begin{aligned}
\frac{\mathrm{d}}{\mathrm{dt}}T(t) &= -\beta T(t)V(t) - \kappa_0 T(t)I(t) + \delta_R R(t)\\
\frac{\mathrm{d}}{\mathrm{dt}}R(t) &= \kappa_0 T(t)I(t) - \delta_R R(t)\\
\frac{\mathrm{d}}{\mathrm{dt}}I(t) &= \beta T(t)V(t) - \left(\delta_I + \kappa_1 D_1(t) + \kappa_2 \mathbb{I}_{t>\tau_A}\right)I(t)\\
\frac{\mathrm{d}}{\mathrm{dt}}V(t) &= pI(t) - cV(t)\\
\frac{\mathrm{d}}{\mathrm{dt}}J(t) &= \nu \frac{D_1(t)^m}{D_1(t)^m + K^m} - \left(\delta_J + \kappa_1 D_1(t)\right)J(t)\\
\frac{\mathrm{d}}{\mathrm{dt}}D_0(t) &= \lambda\left(D_{00} - D_0(t)\right) - \sigma\left(J(t) + I(t)\right)D_0(t)\\
\frac{\mathrm{d}}{\mathrm{dt}}D_1(t) &= \sigma\left(J(t) + I(t)\right)D_0(t) - \delta_D D_1(t).
\end{aligned}\right\} \qquad \text{Eq.(1)}$$

We begin by giving explicit expressions for the possible representations of a resolved infection in the full model (1), we next show that solutions the mathematical model (1) evolving from non-negative initial conditions remain non-negative and bounded, we then determine a positively invariant subset of $\mathbb{R}^7$ and use LaSalle's Invariance Principle to show that the asymptotic behavior of the system is determined by a reduced model system for $J, D_0$ and $D_1$. Finally, we study this reduced system to characterise the local stability of the resolved infection steady state, demonstrate the existence of the hyper inflamed equilibrium, and perform numerical bifurcation analysis.

### Model dynamics of resolved infection

There is an infection free, or resolved, equilibrium solution corresponding to $(T_0, 0,0,0,0, D_{00}, 0)$. This equilibrium is a solution of (3) for any $T_0 \in \mathbb{R}$. It follows that model in Eq. (1) does not admit isolated infection free equilibria, which complicates bifurcation analysis. However, we are primarily interested in the asymptotic dynamics following infection and Eq. (1) has a limited number of target cells $T$. It is then reasonable to expect that all infections not leading to death must eventually resolve and any long-term solutions of Eq. (1) must have $V(t^*)$ and $I(t^*)$ approaching $0$ for $t^*$ large enough. Indeed, any equilibrium solution must satisfy

$$\frac{\mathrm{d}}{\mathrm{dt}}[T(t) + R(t)] = 0,$$

we immediately see that

$$\beta T^* V^* = 0.$$

It follows that $V^* = 0$ or $T^* = 0$. In either case, since $\beta V^* T^* = 0$, the equilibrium condition for $I^*$ becomes

$$0 = -\left(\delta_I + \kappa_1 D_1^* + \kappa_2 \mathbb{I}_{t>\tau_A}\right) I^*.$$

Assuming for now that $D_1^* \geq 0$ (we prove this is the case in Lemma 2), the only possible equilibrium value is $I^* = 0$ as $\delta_I, \kappa_1$, and $\kappa_2$ are strictly positive. The equilibrium condition for $V$ then becomes

$$0 = -cV^*.$$

Therefore, we must have $V^* = 0$ independently of the value of $T^*$. The same argument implies that $I^* = 0$. Thus, informally, for a given $T(0)$ and $R(0)$, the subsystem $T, R, I, V$ has only the trivial equilibrium solution corresponding to the 1-dimensional compact set parametrized by $T^*$

$$L = \{(T^*, 0,0,0) \in \mathbb{R}^4_{\geq 0} | T^* \in [0, T(0) + R(0)]\},$$

where $T^* \leq T(0) + R(0)$ since

$$\frac{\mathrm{d}}{\mathrm{dt}}[T(t) + R(t)] \leq 0$$

and we are assuming that $R \geq 0$ (we show this is the case in Lemma 2).

Then, when considering the long-term behavior of Eq. (1), it is intuitive to consider only the remaining elements of the system. Essentially, the equilibrium values of the subsystem for $T, R, I, V$ must belong to the compact set $L$. Further, since the dynamics of $T$ are contracting and the viral dynamics model is target cell limited, it is natural to expect that all trajectories of $T, R, I, V$ will approach $L$. We formalize this expectation in the following section using LaSalle's Invariance Principle. Accordingly, when considering the long-term behavior of (1), we only consider the subsystem corresponding to $(D_0, D_1, J)$.

#### Model reduction for bifurcation analysis

To show that all long-term trajectories of (1) approach the 1-dimensional compact set $L$ for arbitrary $T^*$, we first recall LaSalle's Invariance Principle

**Theorem 1** (LaSalle's Invariance Principle). *Let $\Sigma$ be a compact positively invariant set with respect to the dynamical system*

$$\frac{\mathrm{d}}{\mathrm{dt}} x(t) = f(x), \quad x(0) = x_0. \qquad \textit{Eq. (2)}$$

*Assume that $\mathcal{V}$ is a negative semi-definite $C_1$ function for all $x \in \Sigma$, let $\Theta \subset \Sigma$ be the set of points such that $\dot{\mathcal{V}}(x) = 0$ for all $x \in \Theta$, and define $\Psi$ as the largest invariant set contained in $\Theta$. Then, all solutions $x(t)$ of Eq. (2) approach $\Psi$ as $t \to \infty$.*

First, we consider the long-time model behavior with $t > \tau_A$ and so include an adaptive immune response in the differential equation for $I(t)$ which becomes

$$\frac{\mathrm{d}}{\mathrm{dt}} I(t) \quad = \beta T(t) V(t) - (\delta_I + \kappa_1 D_1(t) + \kappa_2) I(t).$$

After making this change, Eq. (1) is non-autonomous and takes the place of $f(x)$ in Eq. (2). Therefore, to apply LaSalle's Invariance Principle, we must identify a positively invariant compact set. Given the biological interpretation of Eq. (1), the natural candidate is a bounded subset of the positive octant of $\mathbb{R}^7$. To do this, we begin by showing that trajectories of the full system Eq. (1) evolving from non-negative initial conditions remain non-negative.

**Lemma 2**. *Assume that all parameters of Eq. (1) are strictly positive and let the vector of the initial conditions* $[T(0), R(0), I(0), V(0), J(0), D_0(0), D_1(0)]$ *be component-wise non-negative. Then, solutions of Eq. (1) remain component-wise non-negative for all time* $t > 0$.

*Proof.* We begin by considering the equations for the total number of immune cells. Assume for contradiction that $t_{D_0}$ is the first time such that $D_0(t_{D_0}) = 0$. Then,

$$\frac{\mathrm{d}}{\mathrm{dt}} D_0(t)\Big|_{t=t_{D_0}} = \lambda D_{00} > 0,$$

so $D_0(t)$ is strictly increasing at $t_{D_0}$. Now, if $t_{D_0} = 0$, then $D_0$ becomes strictly positive immediately, while if $t_{D_0} > 0$, then $D_0$ must be non-increasing there, which is a contradiction. Therefore, $D_0(t) > 0$ for all $t > 0$. We now consider the sum $[D_0(t) + D_1(t)]$ and note that

$$\frac{\mathrm{d}}{\mathrm{dt}}[D_0(t) + D_1(t)] \quad = \lambda D_{00} - \lambda D_0(t) - \delta_D D_1(t) < \lambda D_{00} - \min[\lambda, \delta_D]\big(D_0(t) + D_1(t)\big)$$

so the sum must satisfy

$$D_0(t) + D_1(t) \leq \frac{\lambda D_{00}}{\min[\lambda, \delta_D]}.$$

We can thus bound

$$D_1(t) < \max\left[D_1(0), \frac{\lambda D_{00}}{\min[\lambda, \delta_D]}\right] = D_1^{max}.$$

With these estimates in hand, we consider the differential equations for $T(t)$ and $R(t)$. It is simple to see that $R_0 + T_0 = 0$ implies $T(t) + R(t) = 0$ for all time. Furthermore, if $I(0) = V(0) = 0$, then $I(t) = V(t) = 0$ satisfies the differential equations for $I(t)$ and $V(t)$ for all time and imply that the sum $R(t) + T(t)$ is constant. In the infection free case (i.e $I(0) = V(0) = 0$), it follows that

$$R(t) = R_0 \exp[-\delta_R t] \geq 0,$$

and

$$T(t) = T_0 + R_0 \exp[-\delta_R t] \geq 0,$$

which establishes $T(t) \geq 0$ and $R(t) \geq 0$ for $I(0) = V(0) = 0$. We therefore consider $I(0) + V(0) > 0$ and begin with simplest case where $T_0 + R_0 = 0$. With no target cells, it follows that

$$\frac{\mathrm{d}}{\mathrm{dt}} I(t) = -\left(\delta_I + \kappa_1 D_1(t) + \kappa_2 \mathbb{I}_{t>\tau_A}\right) I(t) > -\left(\delta_I + \kappa_1 D_1^{max} + \kappa_2 \mathbb{I}_{t>\tau_A}\right) I(t)$$

and Gronwall's inequality gives

$$I(t) \geq I_0 \exp\left[\left(\delta_I + \kappa_1 D_1^{max} + \kappa_2 \mathbb{I}_{t>\tau_A}\right) t\right] \geq 0.$$

From this estimate, we see

$$\frac{\mathrm{d}}{\mathrm{dt}} V(t) = pI(t) - cV(t) \geq -cV(t)$$

so Gronwall's inequality again demonstrates that $V(t) \geq V_0 \exp(-ct) \geq 0$. This exponential decay is precisely what we expect in the biologically unrealistic target cell free case.

This leaves the case where $T_0 + R_0 > 0$ and $I(0) + V(0) > 0$. We consider three cases:

**Case 1** If $R(0) > 0$ and $T(0) = 0$, we immediately obtain

$$\frac{\mathrm{d}}{\mathrm{dt}} T(t)|_{t=0} = \delta_R R(t) > 0,$$

so $T(t)$ becomes positive immediately while $R(t)$ is strictly positive in a neighbourhood of $t = 0$.

**Case 2** If $T(0) > 0$ and $R(0) = 0$. Either $I(0) > 0$ or $V(0) > 0$ so $I(t)$ becomes positive immediately. In either case,

$$\frac{\mathrm{d}}{\mathrm{dt}} R(t)|_{t=0} = \kappa_0 T(t) I(t) > 0,$$

and so $R(t)$ becomes positive immediately while $T(t)$ is strictly positive in a neighbourhood of $t = 0$.

**Case 3** If both $T(0) > 0$ and $R(0) > 0$, then we have

$$\frac{\mathrm{d}}{\mathrm{dt}} T(t) > -\big(\beta V(t) + \kappa_0 I(t)\big) T(t)$$
$$\frac{\mathrm{d}}{\mathrm{dt}} R(t) > -\delta_R R(t)$$

so Gronwall's inequality implies these quantities decay at most exponentially and thus remain strictly positive. The above arguments show that we can consider $T(0) > 0$ in what follows. We now turn to the differential equations for $I(t)$ and $V(t)$ with $I(0) + V(0) > 0$ and the following three distinct cases **Case I** $V(0) = 0$ and $I(0) > 0$ so we see

$$\frac{\mathrm{d}}{\mathrm{dt}} V(t)|_{t=0} = pI(t) > 0$$

so $V(t)$ becomes strictly positive immediately while $I(t)$ remains strictly positive in a neighbourhood of $t = 0$.

**Case II** $I(0) = 0$ and $V(0) > 0$ so we see

$$\frac{\mathrm{d}}{\mathrm{dt}} I(t)|_{t=0} = \beta T(t) V(t) > 0$$

so $I(t)$ becomes strictly positive immediately while $V(t)$ remains strictly positive in a neighbourhood of $t = 0$.

**Case III** $I(0) > 0$ and $V(0) > 0$. Accordingly, $I(t)$ and $V(t)$ remain strictly positive in a neighbourhood of $t = 0$. While they are non-negative, we compute

$$\begin{aligned}\frac{\mathrm{d}}{\mathrm{dt}}[I(t) + V(t)] &= \beta V(t)T(t) - \left(\delta_I + \kappa_1 D_1(t) + \kappa_2 \mathbb{I}_{t>\tau_A}\right)I(t) + pI(t) - cV(t) \\ &\geq -\left(\delta_I + \kappa_1 D_1(t) + \kappa_2 \mathbb{I}_{t>\tau_A}\right)I(t) - cV(t) \\ &\geq -\max\left[\left(\delta_I + \kappa_1 D_1^{max} + \kappa_2 \mathbb{I}_{t>\tau_A}\right), c\right]\left(I(t) + V(t)\right),\end{aligned}$$

so we once again use Gronwall's inequality to see

$$I(t) + V(t) \geq \left(I(0) + V(0)\right)\exp\left(-\max\left[\left(\delta_I + \kappa_1 D_1^{max} + \kappa_2 \mathbb{I}_{t>\tau_A}\right), c\right]\right) > 0.$$

Thus, if there is a time $t_I$ such that $I(t_I) = 0$, then we argue as in **Case I** Conversely, if there is a time $t_v$ where $V(t_v) = 0$, then $I(t_v) > 0$ and we argue as in **Case II.** Therefore, no such $t_I$ or $t_v$ can exist and we conclude that $I(t)$ and $V(t)$ remain strictly positive for $t > 0$. It follows that $T, R, I$ and $V$ remain strictly non-negative.

Now, we proceed to the remaining equations for $J$ and $D_1$. In the infection free case, if $J(0) + D_1(0) = 0$, then $J(t) = D_1(t) = 0$ solves these two equations. Further, since $I(t) \geq 0$ and $D_0(t) \geq 0$, it follows that

$$\frac{\mathrm{d}}{\mathrm{dt}} D_1(t) \geq \sigma J(t) D_0(t) - \delta_D D_1(t).$$

where we can use Gronwall's inequality to bound $D_1(t)$ by the solution of the above differential equation. Now, assume that $J(0) + D_1(0) > 0$ and consider three cases:

**Case A** Assume $J(0) > 0$ and $D_1(0) = 0$ so we see

$$\frac{\mathrm{d}}{\mathrm{dt}}D_1(t)|_{t=0} \geq \sigma\big(J(t)\big)D_0(t) > 0$$

so $D_1$ becomes strictly positive immediately while $J(t)$ remains strictly positive.

**Case B** Assume that $D_1(0) > 0$ and $J_1(0) = 0$ so we have

$$\frac{\mathrm{d}}{\mathrm{dt}}J(t)|_{t=0} = \nu\frac{D_1(t)^m}{D_1(t)^m + K^m} > 0$$

so $J$ becomes strictly positive immediately while $D_1(t)$ remains strictly positive.

**Case C** Finally, assume that both $J(0)$ and $D_1(0)$ are strictly positive, so $J$ and $D_1$ remain positive in a neighbourhood of $t = 0$. Now, inserting the upper bound for $D_1$ into the differential equation for $J(t)$ gives

$$\frac{\mathrm{d}}{\mathrm{dt}}J(t) \quad = \nu\frac{D_1(t)^m}{D_1(t)^m + K^m} - \big(\delta_J + \kappa_1 D_1^{max}\big)J(t) > -\big(\delta_J + \kappa_1 D_1^{max}\big)J(t),$$

so Gronwall's inequality implies that $J(t) > J(0)\mathrm{exp}\big[-\big(\delta_J + \kappa_1 D_1^{max}\big)t\big] > 0$. This finally implies that

$$\frac{\mathrm{d}}{\mathrm{dt}}D_1(t) > -\delta_D D_1(t),$$

and Gronwall's inequality once again gives $D_1(t) > D_1(0)\mathrm{exp}[-\delta_D t] > 0$. □

We have therefore shown that the non-negative quadrant is positively invariant under Eq. (1). However, the non-negative quadrant is an unbounded subset of $\mathbb{R}^7$, and therefore non-compact, while we need a compact positively invariant set to apply LaSalle's principle. We now show that trajectories of the full system in Eq. (1) are bounded and thus define a positively invariant compact subset of $\mathbb{R}^7$.

**Lemma 3**. *Assume that parameters of (1) are strictly positive and let the vector of the initial conditions* $[T(0), R(0), I(0), V(0), J(0), D_0(0), D_1(0)]$ *be component-wise non-negative. Then, solutions of (1) are bounded above for all time* $t > 0$.

*Proof.* In the proof of Lemma 2, we have already computed an upper bound for $D_1(t)$. Using a similar technique, it follows that

$$D_0(t) + D_1(t) \leq \frac{\lambda D_{00}}{\min[\lambda, \delta_D]},$$

and since $D_1(t) \geq 0$ and $D_0(t) \geq 0$, we conclude

$$D_0(t) < \max\left[D_0(0)\frac{\lambda D_{00}}{\min[\lambda, \delta_D]}\right].$$

Now, consider the differential equation for the sum $T(t) + R(t)$ and see that

$$R(t) + T(t) \leq (R_0 + T_0)\exp\left[-\int_0^t \beta\, V(s)\mathrm{d}s\right] \leq (R_0 + T_0) = T^{max}.$$

as both $R_0$ and $T_0$ are non-negative, $T^{max}$ is an upper bound for each component of the sum.

We now consider the differential equation for $J(t)$

$$\begin{aligned}\frac{\mathrm{d}}{\mathrm{dt}}J(t) &= \nu\frac{D_1(t)^m}{D_1(t)^m + K^m} - \left(\delta_J + \kappa_1 D_1(t)\right)J(t)\\ &\leq \nu - \delta_J J(t)\end{aligned}$$

where the inequality follows from $D_1(t) \geq 0$. It thus follows that, if $J(t) > \frac{\nu}{\delta_J}$, then $J(t)$ is strictly decreasing. It follows that

$$J(t) \leq \max\left[J(0), \frac{\nu}{\delta_J}\right].$$

A simple computation shows that

$$\frac{\mathrm{d}}{\mathrm{dt}}[T(t)+I(t)+R(t)] \leq 0.$$

Therefore, $T(t)+I(t)+R(t) \leq T_0+I_0+R_0$ which implies that $I(t) \leq T_0+I_0+R_0 = I^{max}$. It follows from the differential equation for $V(t)$ that $V(t)$ is strictly decreasing if $V(t) < \frac{pI^{max}}{c}$, which implies

$$V(t) \leq \max\left[V(0), \frac{pI^{max}}{c}\right].$$

□

Combining Lemma 2 and Lemma 3 immediately gives

**Theorem 4**. *Assume that all parameters of Eq. (1) are strictly positive and let the vector of the initial conditions* $[T(0), I(0), V(0), J(0), D_0(0), D_1(0)]$ *be component-wise non-negative. Further, define*

$$T^{max} = T(0)+R(0), I^{max} = T^{max}+I(0), \quad V^{max} = \max\left[V(0), \frac{pI^{max}}{c}\right],$$
$$D_0^{max} = \max\left[D_0(0), \frac{\lambda D_{00}}{\min[\lambda, \delta_D]}\right], D_1^{max} = \max\left[D_1(0), \frac{\lambda D_{00}}{\min[\lambda, \delta_D]}\right], \quad \text{and} \quad J^{max} = \max\left[J(0), \frac{\nu}{\delta_J}\right].$$

*Then, the set*

$$\begin{aligned} E \quad &= \{(T, R, I, V, D_0, D_1, J) \in \mathbb{R}^6 | \\ &T \in [0, T^{max}], R \in [0, R^{max}], I \in [0, I^{max}], V \in [0, V^{max}], \\ &D_0 \in [0, D_0^{max}], D_1 \in [0, D_1^{max}], J \in [0, J^{max}] \} \end{aligned}$$

*is a compact positively invariant set under the flow of (1).*

We now consider the collection of non-isolated equilibria for the subsystem $T(t), R(t), I(t), V(t)$ in $E$

$M = \{(T, 0,0,0, D_0, D_1, J) \in \mathbb{R}^7 | T \in [0, T^{max})], R \in [0, R^{max}],$

$$D_0 \in [0, D_0^{max}], D_1 \in [0, D_1^{max}], J \in [0, J^{max}]\}.$$

It is clear that $M$ is compact and positively invariant under trajectories of (1), and that $M \subset E$. Now, Eq. (1) defines a dynamical system, the set $E$ is a compact positively invariant set under (1), , so to apply LaSalle's invariance principle, we need only define a suitable $\mathcal{V}$. First, fix $x > 0$, set

$$\varepsilon = \frac{\delta_I + \kappa_2/2}{\kappa_0}$$

and take

$$y = 1 + x + \frac{\varepsilon}{T^{max}}.$$

Now, we define

$$\mathcal{V}(T, R, I, V, D_0, D_1, J) = (1 + x)T + yR + I + \frac{(\delta_I + \kappa_2 - \kappa_0\varepsilon)}{p}V.$$

It follows that

$$\begin{aligned}
\dot{\mathcal{V}} \quad &= (1 + x)\frac{\mathrm{d}}{\mathrm{dt}}T(t) + y\frac{\mathrm{d}}{\mathrm{dt}}R(t) + \frac{\mathrm{d}}{\mathrm{dt}}I(t) + \frac{(\delta_I + \kappa_2 - \kappa_0\varepsilon)}{p}\frac{\mathrm{d}}{\mathrm{dt}}V(t) \\
&= -(1 + x)\beta T(t)V(t) - (1 + x - y)\kappa_0 I(t)T(t) + (1 + x - y)\delta_R R(t) + \beta T(t)V(t) \\
&\quad - (\delta_I + \kappa_1 D_1(t) + \kappa_2)I(t) + (\delta_I + \kappa_2)I(t) - \frac{c(\delta_I + \kappa_2 - \kappa_0\varepsilon)}{p}V(t) - \kappa_0\varepsilon I(t) \\
&= -x\beta T(t)V(t) - ((1 + x - y)T(t) + \varepsilon)\kappa_0 I(t) + (1 + x - y)\delta_R R(t) - \kappa_1 D_1(t)I(t)
\end{aligned}$$

$$-\frac{c(\delta_I + \kappa_2 - \kappa_0\varepsilon)}{p}V(t).$$

From the choice of $y$, we immediately have $1 + x - y < 0$ and, recalling that $T(t) \le T^{max}$,

$$(1 + x - y)T(t) + \varepsilon = \left(-\frac{T(t)}{T^{max}} + 1\right)\varepsilon \ge 0.$$

Then, the coefficients of the model variables in $\dot{\mathcal{V}}$ are non-positive. Thus, since all model variables are non-negative, it follows that

$$\dot{\mathcal{V}} \leq 0 \quad \text{for all} \quad (T, R, I, V, D_0, D_1, J) \in E.$$

Furthermore, it is simple to see that $\dot{\mathcal{V}}(T, R, I, V, D_0, D_1, J) = 0$ only if $(T, R, I, V, D_0, D_1, J) \in M$. Thus, LaSalle's Invariance Principle implies that trajectories of (1) converge to $M$. We thus conclude that the long-term behavior of the system is independent of the concentration of target cells and the infection sub-system. Therefore, when performing numerical bifurcation analysis of (1), we consider the reduced system

$$\left.\begin{aligned} \frac{\mathrm{d}}{\mathrm{dt}}J(t) &= \nu \frac{D_1(t)^m}{D_1(t)^m + K^m} - \big(\delta_J + \kappa_1 D_1(t)\big)J(t) \\ \frac{\mathrm{d}}{\mathrm{dt}}D_0(t) &= \lambda\big(D_{00} - D_0(t)\big) - \sigma\big(J(t)\big)D_0(t) \\ \frac{\mathrm{d}}{\mathrm{dt}}D_1(t) &= \sigma\big(J(t)\big)D_0(t) - \delta_D D_1(t) \end{aligned}\right\} \qquad (3)$$

which corresponds to the flow of (1) on $M$.

### Bifurcation analysis

We now consider equilibrium solutions of the reduced 3-dimensional system in (3). Equilibrium solutions of Eq. (3) must satisfy

$$D_0^* = \frac{\lambda D_{00}}{\lambda + \sigma J^*}, \quad D_1^* = \frac{\sigma}{\delta_D}\left(\frac{\lambda D_{00} J^*}{\lambda + \sigma J^*}\right).$$

Inserting these expressions into the equilibrium condition for $J^*$ gives

$$\begin{aligned} 0 &= \nu \frac{\left[\frac{\sigma}{\delta_D}\left(\frac{\lambda D_{00} J^*}{\lambda + \sigma J^*}\right)\right]^m}{\left[\frac{\sigma}{\delta_D}\left(\frac{\lambda D_{00} J^*}{\lambda + \sigma J^*}\right)\right]^m + K^m} - \left[\delta_J + \kappa_1 \frac{\sigma}{\delta_D}\left(\frac{\lambda D_{00} J^*}{\lambda + \sigma J^*}\right)\right]J^* \\ &= \nu \frac{\frac{\sigma}{\delta_D}(\lambda D_{00} J^*)^m}{\frac{\sigma}{\delta_D}(\lambda D_{00} J^*)^m + (\lambda + \sigma J^*)^m K^m} - \left[\delta_J + \kappa_1 \frac{\sigma}{\delta_D}\left(\frac{\lambda D_{00} J^*}{\lambda + \sigma J^*}\right)\right]J^* \end{aligned}$$

which, after eliminating a common denominator, becomes

$$0 = \nu \frac{\sigma}{\delta_D}(\lambda D_{00} J^*)^m - \left(\frac{\sigma}{\delta_D}(\lambda D_{00} J^*)^m + (\lambda + \sigma J^*)^m K^m\right)\left[\delta_J(\lambda + \sigma J^*) + \frac{\kappa_1 \sigma \lambda D_{00} J^*}{\delta_D}\right] J^*$$

the trivial solution $J^* = 0$ is indeed an equilibrium point corresponding to a resolved equilibrium with no cellular damage.

**Analysis of the resolved equilibrium**

The reduced system in Eq. (3) admits the trivial equilibrium $\left(J^h, D_0^h, D_1^h\right) = (0, D_{00}, 0)$ corresponding to a resolved infection in the reduced system (3). The Jacobian Matrix of Eq. (3) evaluated at the resolved steady state is given by

$$\mathcal{J}(0, D_{00}, 0) = \begin{bmatrix} -(\delta_J) & 0 & \frac{\nu m}{K^m} \\ -\sigma D_0^h & -\lambda & 0 \\ \sigma D_0^h & 0 & -\delta_D \end{bmatrix}. \quad (4)$$

The corresponding characteristic equation for the eigenvalues $\xi$ is given by

$$F(\xi) = (\lambda + \xi)\left[\delta_J \delta_D - \frac{\nu m}{K^m}\sigma D_0^h + \left(\delta_J + \delta_D\right)\xi + \xi^2\right].$$

The roots of $F$ are therefore

$$\xi_1 = -\lambda, \quad \xi_2 = \frac{-\left(\delta_J + \delta_D\right) + \sqrt{(\delta_J + \delta_D)^2 - 4[\delta_J \delta_D - \frac{\nu m}{K^M}\sigma D_0^h]}}{2},$$

$$\text{and} \quad \xi_3 = \frac{-\left(\delta_J + \delta_D\right) - \sqrt{(\delta_J + \delta_D)^2 - 4[\delta_J \delta_D - \frac{\nu m}{K^M}\sigma D_0^h]}}{2},$$

which immediately leads to

**Theorem 5**. *The resolved equilibrium is locally asymptotically stable if and only if* $\delta_J \delta_D - \frac{\nu m}{K^M} \sigma D_0^h > 0$.

We can rewrite this condition as

$$\left(\frac{\nu m}{K^M \delta_J}\right)\left(\frac{\sigma D_{00}}{\delta_D}\right) < 1,$$

which acts as a reproduction number type threshold, as the first element in the product represents the amount of inflammation caused by a single activated immune cell and the second element is the number of activated immune cells produced by a single DAMP producing cell.

**Non-resolved equilibrium**

From the above analysis, non-trivial equilibria of (3) must satisfy

$$0 = \nu \frac{\sigma \lambda D_{00}}{\delta_D} (\lambda D_{00} J^*)^{m-1} - \left(\frac{\sigma}{\delta_D} (\lambda D_{00} J^*)^m + (\lambda + \sigma J^*)^m K^m\right)\left[\delta_J (\lambda + \sigma J^*) + \frac{\kappa_1 \sigma}{\delta_D} (\lambda D_{00} J^*)\right].$$

In what follows, we assume that $m \in \mathbb{N}$ and define

$$P_{m+1}(J^*) = \nu \frac{\sigma \lambda D_{00}}{\delta_D} (\lambda D_{00} J^*)^{m-1} - \left(\frac{\sigma}{\delta_D} (\lambda D_{00} J^*)^m + (\lambda + \sigma J^*)^m K^m\right)\left[\delta_J (\lambda + \sigma J^*) + \frac{\kappa_1 \sigma}{\delta_D} (\lambda D_{00} J^*)\right].$$

We immediately see that $P_{m+1}(J^*)$ is a $m+1$ degree polynominal in $J^*$ whose roots determine the equilibrium values of $J^*$. We have already shown that $P_{m+1}(0) = 0$ which corresponds to the resolved equilibrium. Biologically realistic non-trivial equilibria must be strictly positive, so not all the $m+1$ roots of $P_{m+1}$ determine realistic equilibrium of Eq. (3). We now analyze $P_{m+1}(J^*)$ to determine the number of biologically realistic equilibria.

First, to simplify notation, we denote

$$f(J^*) = \left(\frac{\sigma}{\delta_D} (\lambda D_{00} J^*)^m + (\lambda + \sigma J^*)^m K^m\right)\left[\delta_J (\lambda + \sigma J^*) + \frac{\kappa_1 \sigma}{\delta_D} (\lambda D_{00} J^*)\right],$$

so that

$$P_{m+1}(J^*) = \nu \frac{\sigma\lambda D_{00}}{\delta_D}(\lambda D_{00} J^*)^{m-1} - f(J^*).$$

We now expand $f(J^*)$ to re-write $P_{m+1}(J^*)$ in canonical polynomial form. First, we obtain

$$\begin{aligned} f(J^*) \quad &= \frac{\sigma^2\delta_J(\lambda D_{00})^m}{\delta_D}(J^*)^{m+1} + \frac{\sigma\delta_J\lambda^{m+1}D_{00}^m}{\delta_D}(J^*)^m + \frac{\kappa_1\sigma^2(\lambda D_{00})^{m+1}}{\delta_D^2}(J^*)^{m+1} \\ &+ K^m\delta_J(\lambda+\sigma J^*)^{m+1} + \frac{K^m\kappa_1\sigma\lambda D_{00}}{\delta_D}J^*(\lambda+\sigma J^*)^m, \end{aligned}$$

which becomes

$$\begin{aligned} f(J^*) \quad &= \left[\frac{\sigma^2\delta_J(\lambda D_{00})^m}{\delta_D} + \frac{\kappa_1\sigma^2(\lambda D_{00})^{m+1}}{\delta_D^2}\right](J^*)^{m+1} + \frac{\sigma\delta_J\lambda^{m+1}D_{00}^m}{\delta_D}(J^*)^m \\ &+ K^m\delta_J\sum_{n=0}^{m+1}\binom{m+1}{n}\lambda^n(\sigma J^*)^{m+1-n} + \frac{K^m\kappa_1\lambda D_{00}}{\delta_D}\sum_{n=0}^{m}\binom{m}{n}\lambda^n(\sigma J^*)^{m+1-n}. \end{aligned}$$

Regrouping terms in $f(J^*)$ yields

$$\begin{aligned} f(J^*) \quad &= \left[\frac{\sigma^2\delta_J(\lambda D_{00})^m}{\delta_D} + \frac{\kappa_1\sigma^2(\lambda D_{00})^{m+1}}{\delta_D^2} + K^m\delta_J\sigma^{m+1} + \frac{K^m\kappa_1\sigma^{m+1}\lambda D_{00}}{\delta_D}\right](J^*)^{m+1} \\ &+ \left[\frac{\sigma\delta_J\lambda^{m+1}D_{00}^m}{\delta_D} + K^m\delta_J\binom{m+1}{m}\lambda\sigma^m + \frac{K^m\kappa_1\lambda D_{00}}{\delta_D}\binom{m}{m-1}\lambda\sigma^m\right](J^*)^m \\ &+ K^m\delta_J\sum_{n=2}^{m+1}\binom{m+1}{n}\lambda^n(\sigma J^*)^{m+1-n} + \frac{K^m\kappa_1\lambda D_{00}}{\delta_D}\sum_{n=2}^{m}\binom{m}{n}\lambda^n(\sigma J^*)^{m+1-n} \end{aligned}$$

which gives

$$\begin{aligned} f(J^*) \quad &= \left[\frac{\sigma^2\delta_J(\lambda D_{00})^m}{\delta_D} + \frac{\kappa_1\sigma^2(\lambda D_{00})^{m+1}}{\delta_D^2} + K^m\delta_J\sigma^{m+1} + \frac{K^m\kappa_1\sigma^{m+1}\lambda D_{00}}{\delta_D}\right](J^*)^{m+1} \\ &+ \left[\frac{\sigma\delta_J\lambda^{m+1}D_{00}^m}{\delta_D} + K^m\delta_J\binom{m+1}{m}\lambda\sigma^m + \frac{K^m\kappa_1\lambda D_{00}}{\delta_D}\binom{m}{m-1}\lambda\sigma^m\right](J^*)^m \\ &+ \sum_{n=2}^{m}\left(K^m\delta_J\binom{m+1}{n} + \frac{K^m\kappa_1\lambda D_{00}}{\delta_D}\binom{m}{n}\right)\lambda^n(\sigma J^*)^{m+1-n} + K^m\delta_J\lambda^{m+1}. \end{aligned}$$

Therefore, we see that

$$\left.\begin{aligned} P_{m+1}(J^*) \quad &= -\left[\frac{\sigma^2\delta_J(\lambda D_{00})^m}{\delta_D} + \frac{\kappa_1\sigma^2(\lambda D_{00})^{m+1}}{\delta_D^2} + K^m\delta_J\sigma^{m+1} + \frac{K^m\kappa_1\sigma^{m+1}\lambda D_{00}}{\delta_D}\right](J^*)^{m+1} \\ &- \left[\frac{\sigma\delta_J\lambda^{m+1}D_{00}^m}{\delta_D} + K^m\delta_J\binom{m+1}{m}\lambda\sigma^m + \frac{K^m\kappa_1\lambda D_{00}}{\delta_D}\binom{m}{m-1}\lambda\sigma^m\right](J^*)^m \\ &+ \left(\nu\frac{\sigma\lambda D_{00}}{\delta_D}(\lambda D_{00})^{m-1} - K^m\delta_J\binom{m+1}{2} - \frac{K^m\kappa_1\lambda D_{00}}{\delta_D}\binom{m}{2}\right)(J^*)^{m-1} \\ &- \sum_{n=3}^{m}\left(K^m\delta_J\binom{m+1}{n} + \frac{K^m\kappa_1\lambda D_{00}}{\delta_D}\binom{m}{n}\right)\lambda^n(\sigma J^*)^{m+1-n} - K^m\delta_J\lambda^{m+1}, \end{aligned}\right\} \qquad (5)$$

or alternatively,

$$P_{m+1}(J^*) = \sum_{n=0}^{m+1} a_n\,(J^*)^n,$$

where the coefficients $a_n$ are given in Eq. (5). Importantly, we note that all the coefficients except $a_{m-1}$ are strictly negative while

$$a_{m-1} = \nu\frac{\sigma\lambda D_{00}}{\delta_D}(\lambda D_{00})^{m-1} - K^m\delta_J\binom{m+1}{2} - \frac{K^m\kappa_1\lambda D_{00}}{\delta_D}\binom{m}{2}$$

can take either sign.

Recalling that we are only interested in strictly positive solutions of $P_{m+1}(J^*) = 0$, corresponding to biologically relevant equilibria, we utilize *Descartes' rule of signs* to determine the number of equilibria solutions. We recall that *Descartes' rule of signs* states that the number of positive roots of a polynomial $g(x) = \sum_{n=0}^{m}\alpha_n\,x^m$ is at most the number of sign changes in the sequence $\{\alpha_n\}$ and that the difference between the number of sign changes and the number of positive roots is always even.

We now show that there are at most two positive equilibria.

**Theorem 6**. *Assume that all parameters of Eq. (3) are strictly positive and* $m \in \mathbb{N}$ *with* $m \geq 2$*. Then, there are at most two non-trivial equilibrium solutions.*

*Proof.* First, assume that $a_{m-1} \leq 0$, so the sequence of coefficients $a_n$ has no sign changes. Then, *Descartes' rule of signs* implies that there are no positive solutions of $P_{m+1}(J^*) = 0$ and thus no non-trivial equilibria solutions.

Now, assume that $a_{m-1} > 0$. Then, the sequence $\{a_n\}$ changes sign between the pairs $a_m$ and $a_{m-1}$ and $a_{m-1}$ and $a_{m-2}$. Therefore, *Descartes' rule of signs* implies that there are either 2 or 0 positive solutions of $P_{m+1}(J^*) = 0$. This establishes the claim. □

Numerical simulation indicates the existence of a stable non-trivial equilibrium that disappears as $\kappa_1$ increases. These numerical observations, combined with the upper bounds on the number of equilibria solutions established in Theorem 6 suggests the presence of a saddle-node bifurcation. Indeed, we note that, by considering $a_{m-1}$ as a function of model parameters, $a_{m-1}$ is monotonically decreasing in $\kappa_1$. Then, simple inspection suggests $\kappa_1$ as a candidate bifurcation parameter as $a_{m-1}(\kappa_1^*) = 0$ for

$$\kappa_1^* = \frac{\nu \frac{\sigma \lambda D_{00}}{\delta_D} (\lambda D_{00})^{m-1} - K^m \delta_J \binom{m+1}{2}}{\frac{K^m \lambda D_{00}}{\delta_D} \binom{m}{2}}.$$

Similar reasoning also suggests performing bifurcation analysis in $\nu$ with

$$\nu^* = \frac{K^m \delta_J \binom{m+1}{2} + \frac{K^m \kappa_1 \lambda D_{00}}{\delta_D} \binom{m}{2}}{\frac{\sigma \lambda D_{00}}{\delta_D} (\lambda D_{00})^{m-1}}.$$

To test these predictions, we performed numerical bifurcation analysis using Matcont [Dhooge et al., 2008]. We identified a Saddle-Node bifurcation in Eq. (3) that is shown in Fig S1. We performed further continuation analysis to interrogate the relationship between $\nu$ and $\kappa_1$ by following the Saddle-Node bifurcation point in Fig S2. Our results indicate that increases in immune activation by damaged cells, $\nu$, can be counterbalanced by increasing immune efficacy of clearing damaged cells $\kappa_1$ to avoid convergence to the hyper inflamed steady state.

**Infection drives the system into the basin of attraction of the hyper inflamed equilibrium**

In the reduced model in Eq. (3), solution trajectories cannot cross the separatrix that appears following the Saddle-Node bifurcation. Consequently, the dynamics of the non-autonomous differential equation are determined by the initial concentration of damaged cells, $J(0)$. However, the infection subsystem $(T, R, I, V)$ acts to induce cellular damage via infection. This infection driven damage can force the system across the separatrix and into the basin of attraction of the hyper inflamed equilibrium. Our analysis thus indicates a potential mechanism by which viral infection can lead to sufficient bystander cell damage to drive the inflammatory response into a self-sustaining positive feedback loop corresponding to convergence to the hyper inflamed equilibria.

**References**


Dhooge, A., Govaerts, W., Kuznetsov, Y. A., Meijer, H. G., and Sautois, B. (2008). New features of the software MatCont for bifurcation analysis of dynamical systems. *Math. Comput. Model. Dyn. Syst.*, 14(2):147–175.

### Supplementary Figures

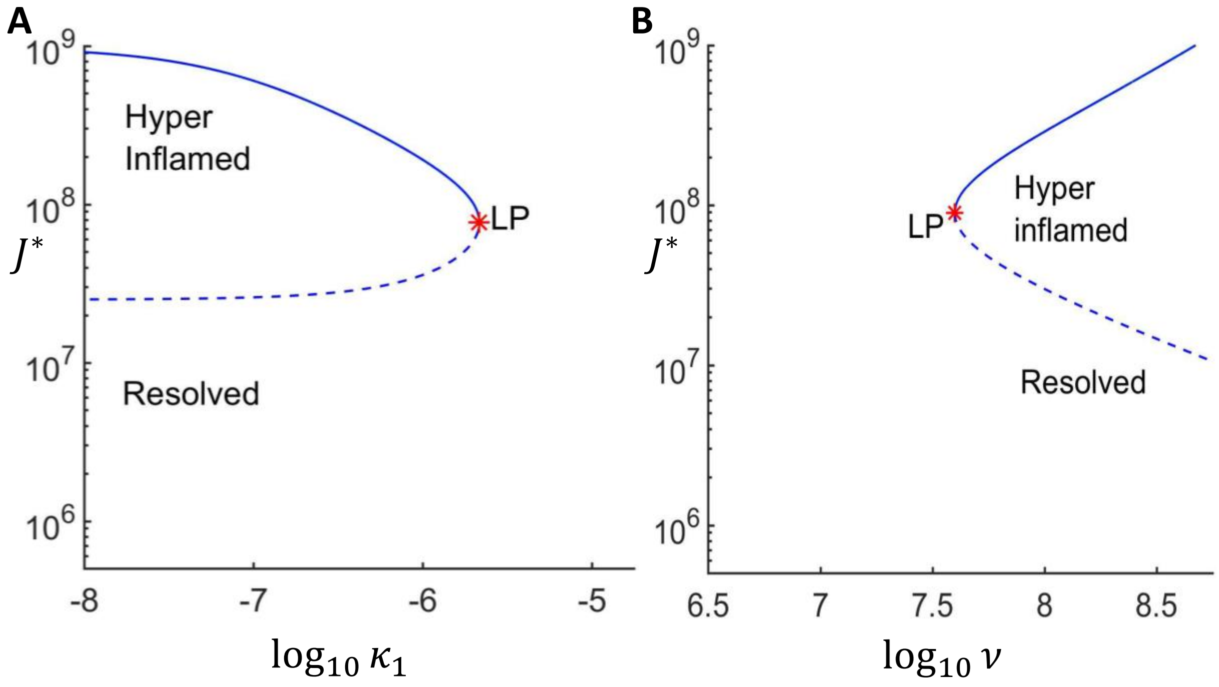


**Figure S1: The hyperinflamed state appears following a saddle node bifurcation.** The non-resolved equilibrium values of $J^*$ as a function of the bifurcation parameter. The saddle-node bifurcation point is denoted by *LP*. The solid branch denotes the locally asymptotically stable hyper inflamed equilibrium while the lower dashed branch of equilibria is unstable. The unstable branch acts as a separatrix between the resolved and hyper inflamed states. Viral infection drives model trajectories across the separatrix and into the basin of attraction of the hyper inflamed steady state. In the simulations, this convergence to the hyper inflamed steady state corresponding to the severe COVID-19 group while simulations that remain in the resolved basin of attraction converge to the resolved steady state. Panel A plots the equilibrium values $J^*$ as a function of $\log_{10} \kappa_1$ while panel B values $J^*$ as a function of $\log_{10} \nu$.

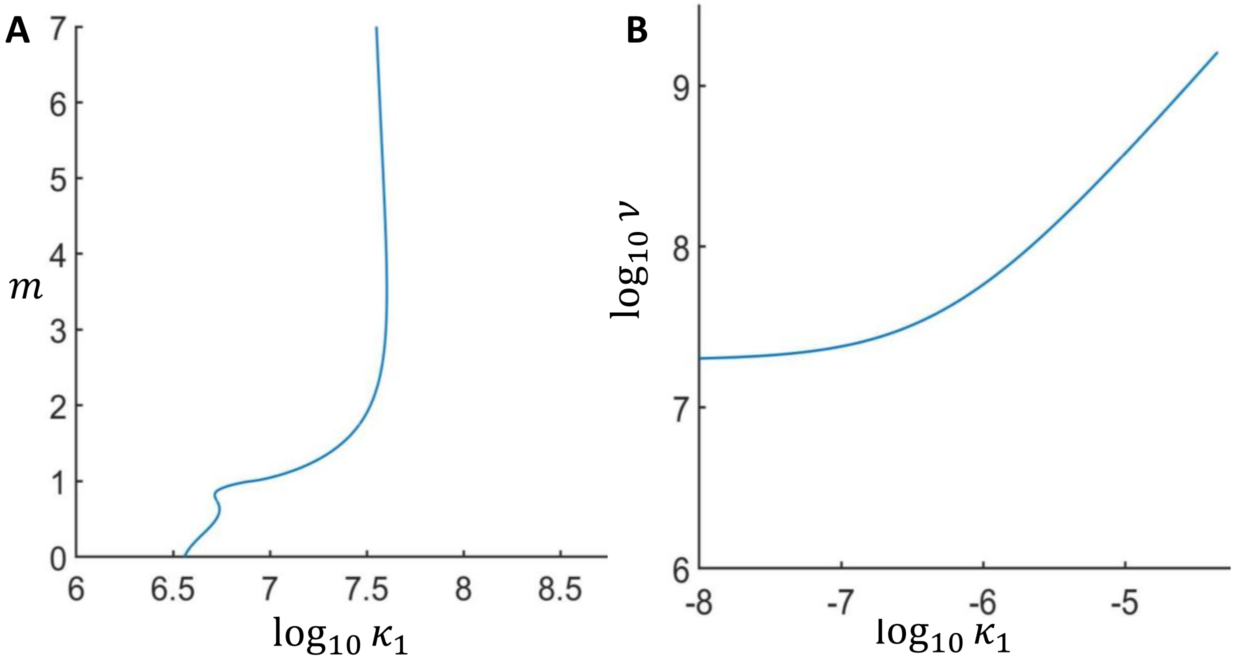


**Figure S2: Continuation of the saddle-node bifurcation in two model parameters.** The curves plot the location of the saddle-node bifurcation as two model parameters vary simultaneously and shows the non-linear relationship between model parameters. Panel A illustrates how decreases in $m$ can be counterbalanced by changes in $\log_{10} \nu$ to allow for the existence of a hyper inflamed state. Panel B shows how increased immune-mediated resolution of inflammation, corresponding to larger values of $\kappa_1$, can be overcome by increased immune-mediated damage, or increased $\nu$, to allow for a saddle node bifurcation and the resulting hyper inflamed state.

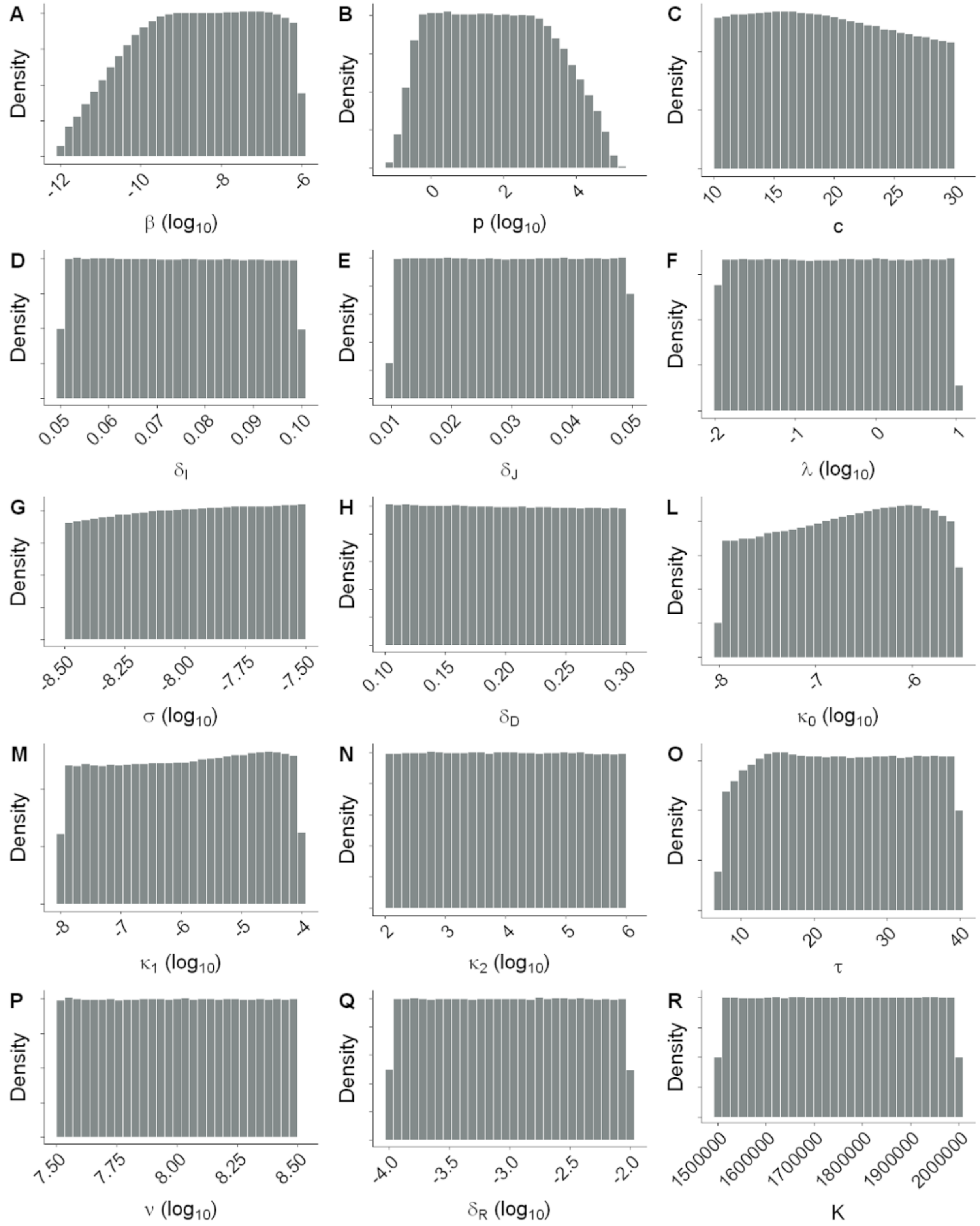


**Figure S3. Distribution of parameters leading to accepted simulations based on conditions i)-iii).**

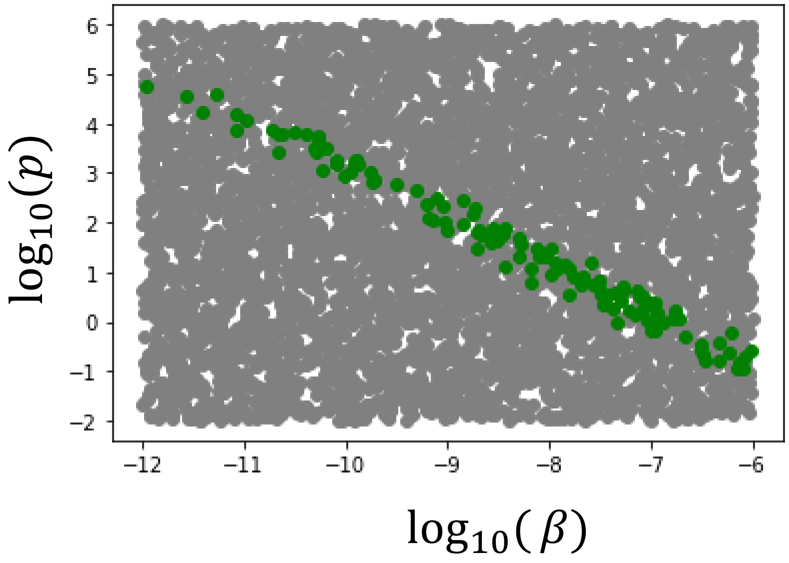


**Figure S4: Linear relationship between $\log_{10}\beta$ and $\log_{10}p$ in the space of acceptable parameter values.** Values leading to acceptance of conditions i)-iii) are represented bv green dots, while those values that lead to unacceptable viral loads are represented by grey dots.

| | $\beta(\log_{10})$ | c | $\delta_I$ | $\delta_J$ | $\kappa_0(\log_{10})$ | $\delta_R(\log_{10})$ | $\kappa_1(\log_{10})$ | $\kappa_2$ | $\nu(\log_{10})$ | K | $\lambda(\log_{10})$ | $\sigma(\log_{10})$ | $\delta_D(\log_{10})$ | $\tau(\log_{10})$ | Score($\log_{10}$) | p($\log_{10}$) |
|---|---|---|---|---|---|---|---|---|---|---|---|---|---|---|---|---|
| $\beta(\log_{10})$ | | -0.05 | 0 | 0 | -0.16 | 0 | -0.05 | 0 | 0 | 0 | 0 | -0.03 | 0 | 0 | 0.2 | -0.99 |
| c | -0.05 | | 0 | 0 | -0.01 | 0 | 0.01 | 0 | 0 | 0 | 0 | 0.01 | 0 | 0.01 | -0.09 | 0.08 |
| $\delta_I$ | 0 | 0 | | 0 | 0 | 0 | 0 | 0 | 0 | 0 | 0 | 0 | 0 | 0 | 0 | 0 |
| $\delta_J$ | 0 | 0 | 0 | | 0 | 0 | 0 | 0 | 0 | 0 | 0 | 0 | 0 | 0 | 0 | 0 |
| $\kappa_0(\log_{10})$ | -0.16 | -0.01 | 0 | 0 | | 0 | -0.05 | 0 | 0 | 0 | 0 | 0.03 | -0.01 | 0.02 | -0.66 | 0.17 |
| $\delta_R(\log_{10})$ | 0 | 0 | 0 | 0 | 0 | | 0 | 0 | 0 | 0 | 0 | 0 | 0 | 0 | 0.01 | 0 |
| $\kappa_1(\log_{10})$ | -0.05 | 0.01 | 0 | 0 | -0.05 | 0 | | 0 | 0 | 0 | 0 | 0.01 | 0 | 0.01 | -0.27 | 0.05 |
| $\kappa_2$ | 0 | 0 | 0 | 0 | 0 | 0 | 0 | | 0 | 0 | 0 | 0 | 0 | 0 | 0 | 0 |
| $\nu(\log_{10})$ | 0 | 0 | 0 | 0 | 0 | 0 | 0 | 0 | | 0 | 0 | 0 | 0 | 0 | 0.12 | 0 |
| K | 0 | 0 | 0 | 0 | 0 | 0 | 0 | 0 | 0 | | 0 | 0 | 0 | 0 | -0.03 | 0 |
| $\lambda(\log_{10})$ | 0 | 0 | 0 | 0 | 0 | 0 | 0 | 0 | 0 | 0 | | 0 | 0 | 0 | 0.18 | 0 |
| $\sigma(\log_{10})$ | -0.03 | 0.01 | 0 | 0 | 0.03 | 0 | 0.01 | 0 | 0 | 0 | 0 | | 0.01 | -0.01 | 0.11 | 0.03 |
| $\delta_D(\log_{10})$ | 0 | 0 | 0 | 0 | -0.01 | 0 | 0 | 0 | 0 | 0 | 0 | 0.01 | | 0 | -0.06 | 0 |
| $\tau(\log_{10})$ | 0 | 0.01 | 0 | 0 | 0.02 | 0 | 0.01 | 0 | 0 | 0 | 0 | -0.01 | 0 | | 0.04 | 0 |
| Score($\log_{10}$) | 0.2 | -0.09 | 0 | 0 | -0.66 | 0.01 | -0.27 | 0 | 0.12 | -0.03 | 0.18 | 0.11 | -0.06 | 0.04 | | -0.16 |
| p($\log_{10}$) | -0.99 | 0.08 | 0 | 0 | 0.17 | 0 | 0.05 | 0 | 0 | 0 | 0 | 0.03 | 0 | 0 | -0.16 | |

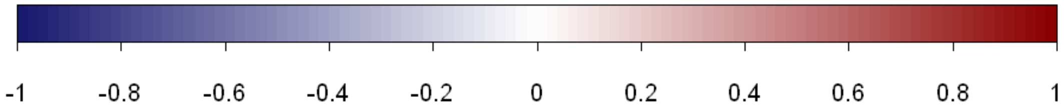


**Figure S5. Correlation matrix between model parameters and between model parameters and Disease Score**

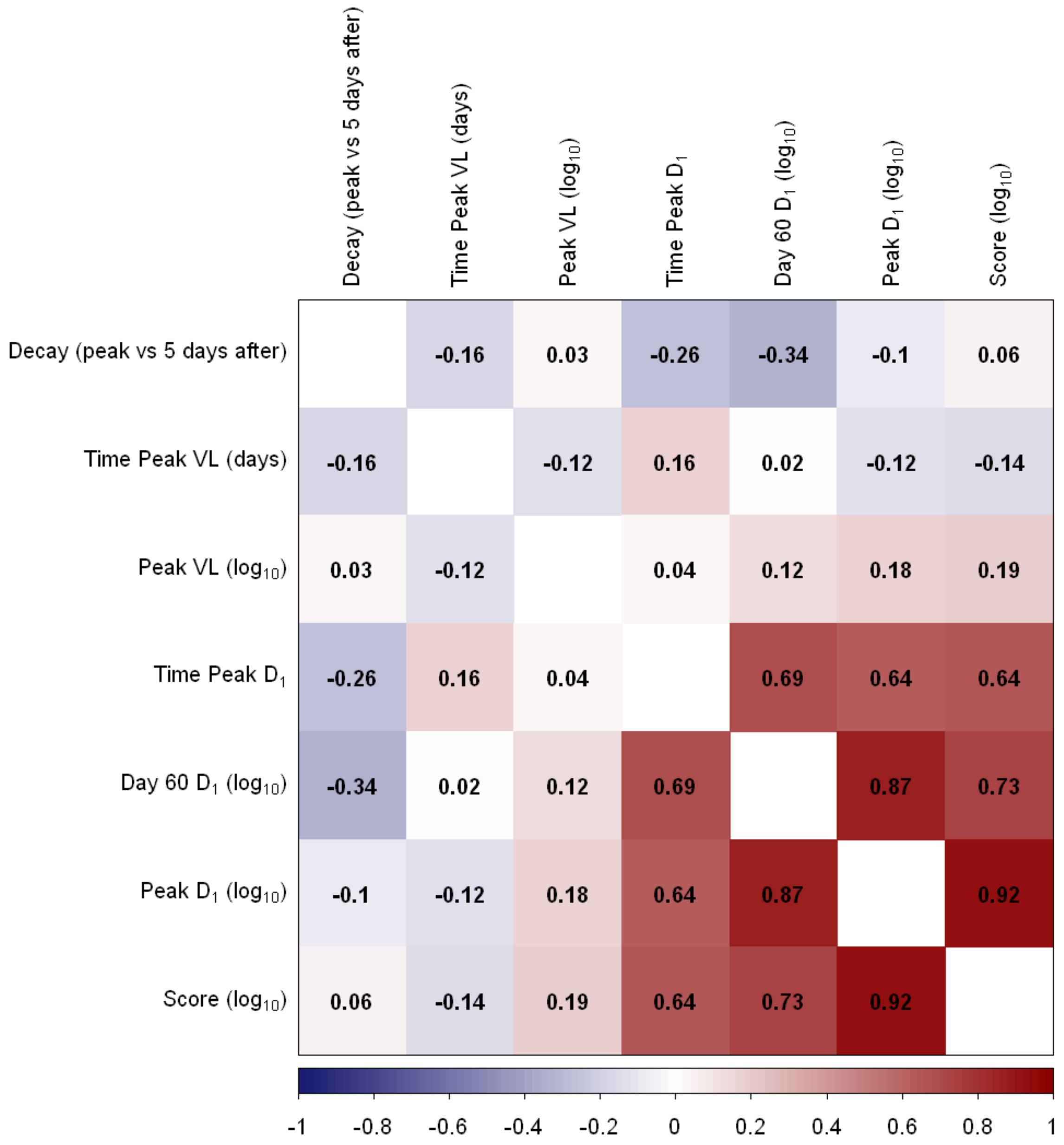


**Figure S6. Correlation matrix between virtual markers and between markers and Disease Score**

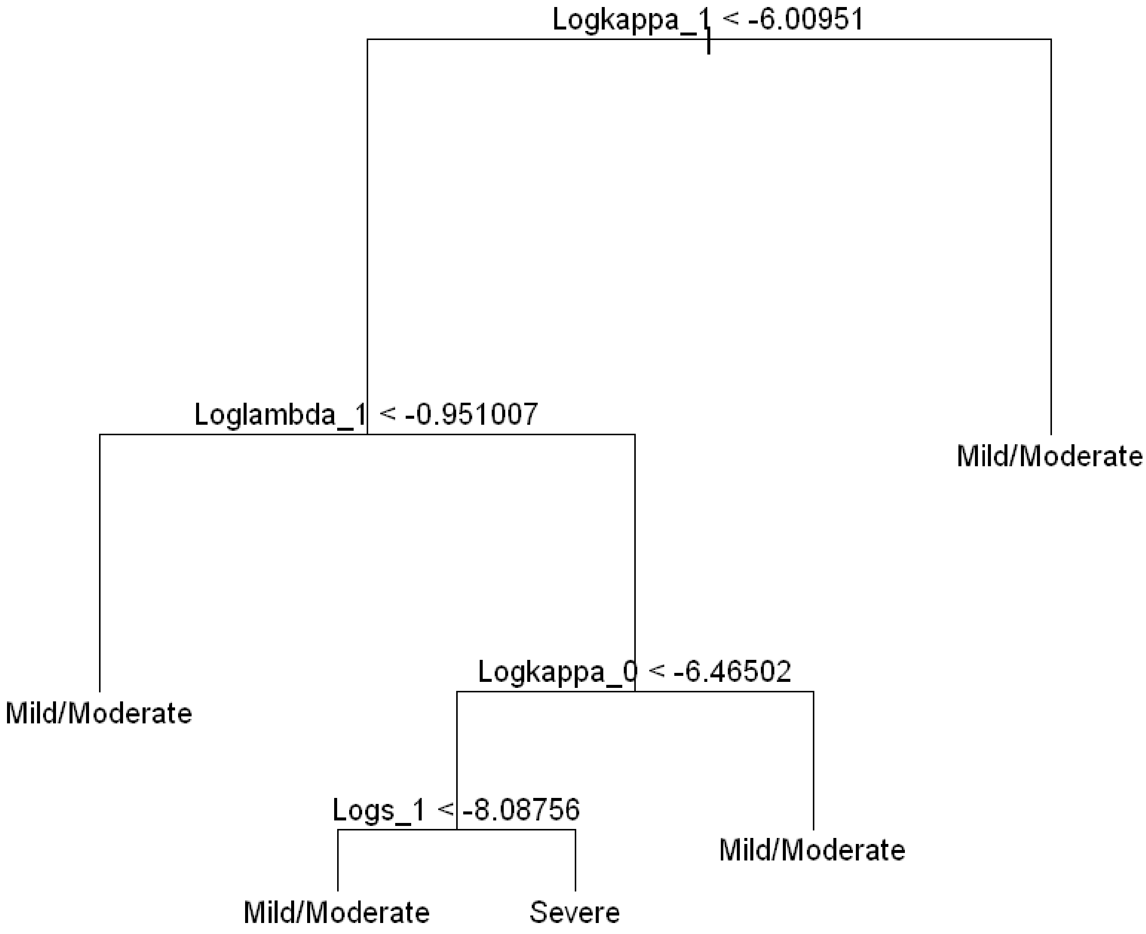


**Figure S7. Complete tree for predicting hyperinflammation from model parameters.** Resolved inflammation is represented by label Mild/Moderate. Hyperinflammation is represented by label Severe.

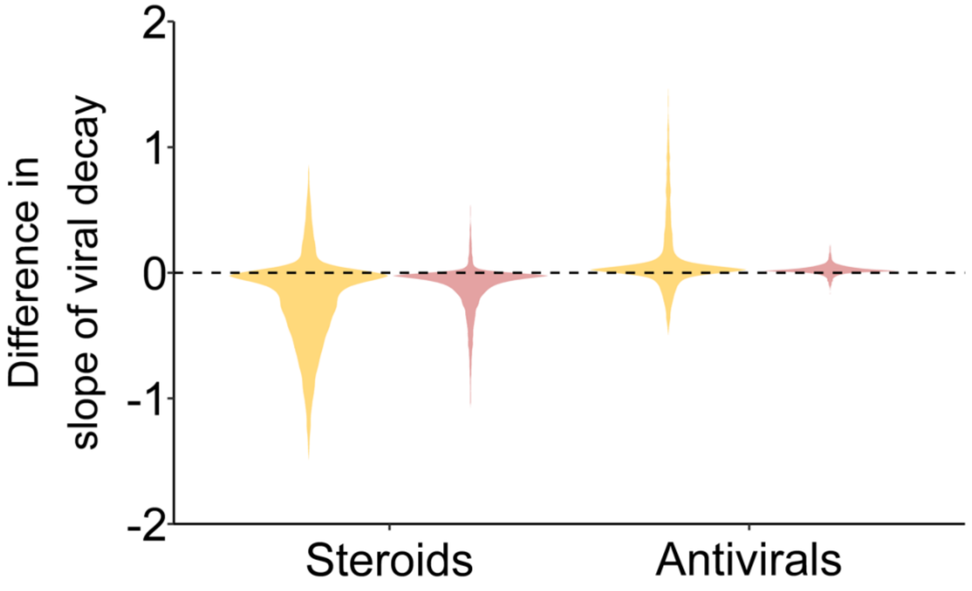


**Figure S8. Violin plots of the effect of virtual treatment on the slope of viral load decay after the peak viral load.** Negative values represent slower viral load decay after treatment administration, while positive values represent faster decay. Note there was no clear difference between a reduction in $\beta$ or $p$ in the simulation of antivirals so this plot applies to both cases. Orange denotes the effect of treatment among individuals who would have resolved inflammation in the absence of treatment, whereas pink denotes the effect of treatment among individuals who would have had hyperinflammation in the absence of treatment.
